# Towards the sustainable elimination of human African trypanosomiasis in Côte d’Ivoire using an integrated approach

**DOI:** 10.1101/2023.02.17.23285863

**Authors:** Dramane Kaba, Mathurin Koffi, Lingué Kouakou, Emmanuel Kouassi N’Gouan, Vincent Djohan, Fabrice Courtin, Martial Kassi N’Djetchi, Bamoro Coulibaly, Guy Pacôme Adingra, Djakaridja Berté, Bi Tra Dieudonné Ta, Minayégninrin Koné, Samuel A Sutherland, Ron E Crump, Ching-I Huang, Jason Madan, Paul R Bessell, Antoine Barreaux, Philippe Solano, Emily H Crowley, Kat S Rock, Vincent Jamonneau

## Abstract

**Background:** Human African trypanosomiasis is a parasitic disease caused by trypanosomes among which *Trypanosoma brucei gambiense* is responsible for a chronic form (gHAT) in West and Central Africa. Its elimination as a public health problem (EPHP) is being achieved. Côte d’Ivoire was one of the first countries to be validated by WHO in 2020 and this was particularly challenging as the country still reported around a hundred cases a year in the early 2000s. This article describes the strategies implemented including a mathematical model to evaluate the reporting results and infer progress towards sustainable elimination.

**Methods:** The control methods used combined both exhaustive and targeted medical surveillance strategies to diagnose and treat cases as well as vector control to reduce the risk of transmission in the most at risk areas. A mechanistic model was used to estimate the number of underlying infections and the probability of elimination of transmission (EoT) between 2000– 2021 in two endemic and two hypo-endemic health districts.

**Results:** Between 2015 and 2019, nine gHAT cases were detected in two health districts in which the number of cases/10,000 inhabitants was far below 1, a necessary condition for validating the EPHP. Modelling estimated a slow but steady decline in transmission across the four health districts, bolstered in the two endemic health districts by the introduction of vector control. The decrease in underlying transmission in all health districts corresponds to a high probability that EoT has already occurred in Côte d’Ivoire.

**Conclusion:** This success was achieved through a multi-stakeholder and multidisciplinary one health approach where research has played a major role in adapting tools and strategies to this large epidemiological transition to a very low prevalence. This integrated approach will need to continue to reach the verification of EoT in Côte d’Ivoire targeted by 2025.

**Author Summary:** Significant efforts to control *Trypanosoma brucei gambiense* human African trypanosomiasis (gHAT) have drastically reduced the prevalence of the disease and elimination of transmission (EoT) is targeted for 2030 by WHO. This reduction was particularly challenging in Côte d’Ivoire as it still faced epidemic episodes in the early 2000s. This large epidemiological transition to very low prevalence necessitated the adaptation and evolution of both medical and vector control strategies described in this article. A mathematical model was also used to retrospectively analyse case reporting results, indicating with high probability that local EoT has already been achieved in the four health districts analysed.

With nine gHAT cases detected in two health districts between 2015 and 2019 and less than one case per 10,000 people per year in all health districts at national level over this five-year period, Côte d’Ivoire received validation by WHO of achievement of the elimination of the disease as a public health problem in 2020. These results combined with the modelling offer encouragement regarding reaching the verification of EoT targeted by 2025 in Côte d’Ivoire on condition of maintaining such multidisciplinary one health approach including research activities to continuously adapt it to the epidemiological transition to zero incidence.

## Introduction

Human African trypanosomiasis (HAT) is a parasitic disease caused by trypanosomes that are transmitted to humans by tsetse [1]. *Trypanosoma brucei gambiense* (*T. b. gambiense*) is responsible for a chronic form of HAT (*gambiense* HAT, gHAT) in West and Central Africa accounting for 97% of all reported HAT cases during 2001–2020 [2]. In comparison, *T. b. rhodesiense* is zoonotic and responsible for an acute form of HAT (*rhodesiense* HAT, rHAT) in East and South Africa. Both forms can be deadly if left untreated [3]. From the 1970s to the 1990s, HAT experienced a phase of emergence/re-emergence that resulted in a significant increase in the number of cases, peaking at 37,385 reported cases recorded in 1998 [4]. The response to increasing cases, which was organised around national programmes of endemic countries dedicated to HAT control supported by the World Health Organization (WHO) and partners, was effective, and the number of cases reported annually fell below 10,000 in 2009 [5]. gHAT was then included in the WHO roadmap for neglected tropical diseases (NTDs) in 2012, with the goal of elimination as a public health problem (EPHP) by 2020 [6] and subsequently elimination of transmission (EoT) to humans by 2030 [2].

Two main global indicators were subsequently defined to monitor the EPHP process: 1) reducing the annual number of cases to fewer than 2,000 per year by 2020; and 2) a 90% reduction in the area at moderate, high or very high risk (the latter defined as an area that reports in excess of one case/10,000 people/year, averaged over a five-year period). The 90% reduction refers to the period 2016–2020 compared to the 2000–2004 reference period. With fewer than 1,000 cases reported each year since 2018 (565 gHAT and 98 rHAT cases were reported in 2020) the first global indicator was met, however a reduction of 120,000 km^2^ (83%) in the moderate and high-risk area meant the global 2020 EPHP target was slightly missed but was thought to be achievable by 2022 [2]. Country-specific validation of EPHP by the WHO is conducted through the compilation of a dossier of data to demonstrate that the indicator for national EPHP has been achieved. This indicator is defined as an average of less than one case per 10,000 people per year over a five-year period in each health district. Togo and Côte d’Ivoire were the first countries to be validated by WHO as having achieved EPHP of gHAT [2].

Achieving this goal was challenging in Côte d’Ivoire as the country reported around a hundred gHAT cases a year in the early 2000s [7], the majority of which were in endemic foci in Western-Central forest areas [8]. Since 2009 fewer than 10 gHAT cases were reported annually with the reduction in transmission and detected case numbers driven by active case detection by mobile teams that screen at-risk populations [7,9]. This large epidemiological transition to very low incidence necessitates the adaptation and evolution of control strategies since there are diminishing returns for the same level of effort. This experience is shared by other elimination or eradication programmes such as polio [10] or Guinea worm [11] where previous rapid reductions in case reporting have now been replaced by extremely low but persistent case detections and new intervention approaches have been required. To overcome this end-game challenge, gHAT strategies in Côte d’Ivoire have now shifted to utilise innovative tools both within the classical “screen, diagnose and treat” algorithm and by use of complementary vector control [12]. This article describes the approach led by the Programme National d’Elimination de la THA (PNETHA) by focusing on the case reporting results obtained during the period 2015–2019 that enabled the validation of EPHP in Côte d’Ivoire. We also used a mathematical modelling approach to quantitatively evaluate these reporting results and infer progress towards the achievement of EoT of gHAT, demonstrating the importance of an integrated, data-driven approach for sustainable elimination.

## Materials and methods

In this section we describe details of the epidemiological context of gHAT in Côte d’Ivoire, the recent activities in intervention and surveillance by PNETHA, and outline how we have utilised mathematical modelling to retrospectively analyse case reporting results. The paper focuses on the 2015–2019 period on which the EPHP was based, but also takes into account the number of cases reported by PNETHA historically (2000–2014) and after the EPHP dossier was submitted (2020–2021) to further demonstrate the impact of the programme through modelling.

### Study area

gHAT is characterised in Côte d’Ivoire, as it is throughout Africa, as a focal disease with hotspots of infection [13,14]. Although the focus boundaries are difficult to precisely define, the foci were considered to be epidemiological units until 2014 (S1 Figure). Subsequently, health districts (HD) have been used as the epidemiological units of analysis as a requirement for reporting to WHO for the EPHP. The number of reported cases per HD in Côte d’Ivoire between 2000 and 2014 is given in S1 Table. In 2015 HDs in Côte d’Ivoire can be categorised into four distinct groups (Fig 1):

**Fig. 1.**
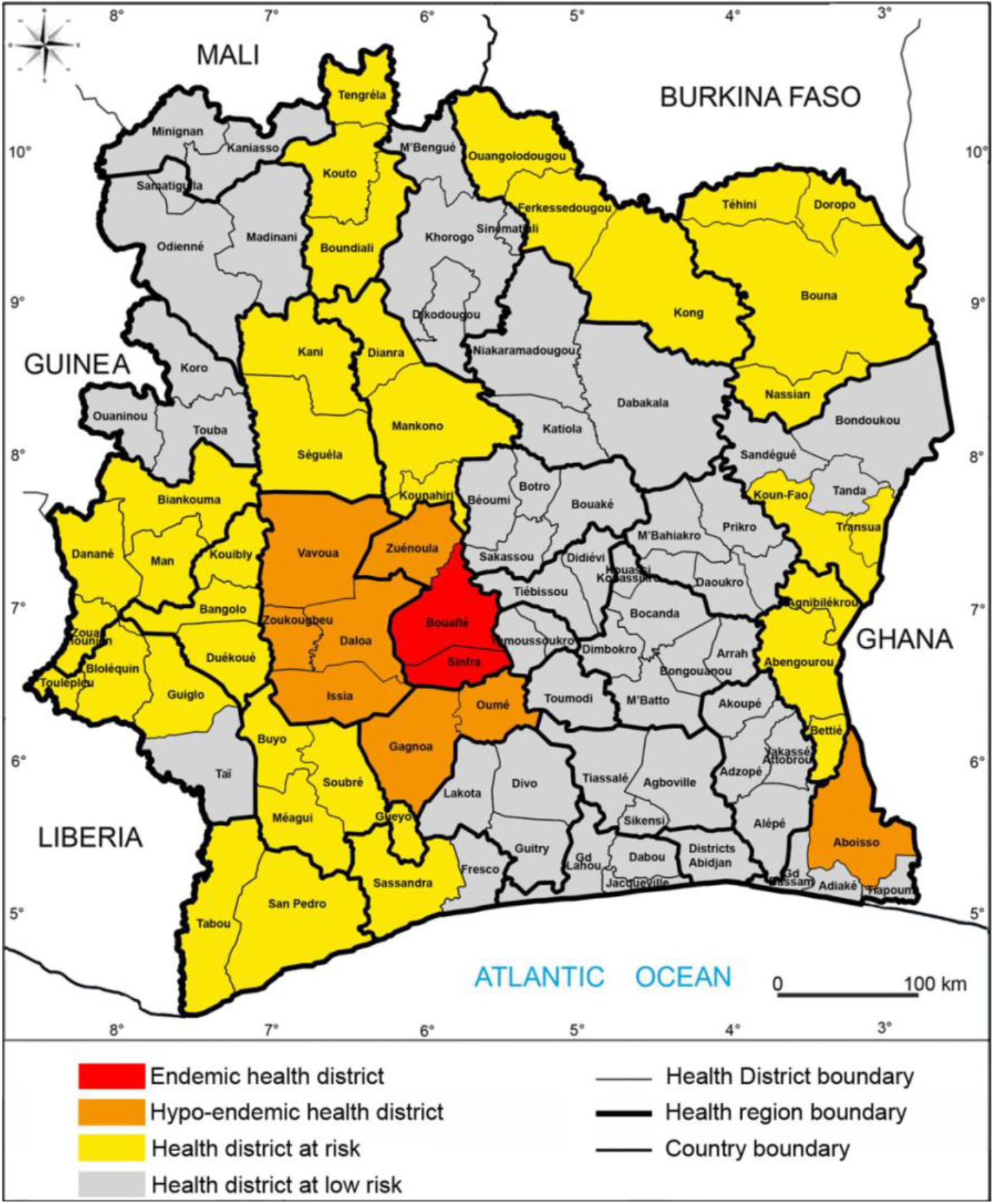
Study areas and gHAT epidemiological status since 2015. Source: Institut Pierre Richet, BNEDT/Openstreetmap.org

1. The *endemic HDs* of Bouaflé (including the subprefecture of Bonon) and Sinfra which reported cases each year during the period 2000–2010.
2. The *hypo-endemic HDs* of Aboisso, Daloa, Issia, Gagnoa, Oumé, Vavoua, Zuénoula and Zoukougbeu which have also reported cases between 2000–2010 but not every year.
3. The *HDs at risk* in which cases have been reported prior to but not since 2000.
4. The *HDs at low risk* where there has been very little (often unproven) or no past reporting of gHAT before 2000 but where the presence of tsetse is likely (allowing for potential transmission).

Like most other neglected tropical diseases (NTDs), gHAT is not vaccine preventable, so control and elimination activities have been focused on two key parts of the transmission cycle:

1. Treatment of infected individuals which acts both to prevent disease mortality but also to reduce the time people spend infected and infectious to tsetse vectors.
2. Targeting the tsetse vector with the goal of reducing the number of transmission events.

Unlike several other NTDs, the treatment pathway for gHAT currently requires confirmation of infection prior to treatment, so mass drug administration or “screen-and-treat” strategies cannot currently be used. The range of medical and vector interventions routinely recommended and deployed to tackle gHAT are presented elsewhere [3]. However, the gHAT control strategy is variable from region to region and so here we outline the different screening, diagnostic and vector control approaches that have formed part of the gHAT elimination strategy in Côte d’Ivoire during the 2015–2019 period.

### Active screening and targeted active screening

The first step for active screening carried out by the mobile teams involved informing the administrative and customary authorities and raising awareness among the populations targeted for screening – typically active screening has aimed to recruit as many people as possible in villages identified for screening. Diagnosis was made based on the decision algorithm shown in Fig 2a. The card agglutination test for trypanosomiasis (CATT) [15] on whole blood collected by finger puncture (CATTb) was carried out in all the people who presented themselves to the mobile team during the screening activity. In the case of a positive result with CATTb, a sample was taken from the bend of the elbow (5ml of heparinized blood) to perform the CATT on plasma dilution (CATTp). “Serosuspects” were defined to be individuals with a positive CATTp at a dilution of at least ¼, and they underwent parasitological examination: miniature anion-exchange centrifugation technique with buffy coat (mAECT BC, [16]) and microscopic examination between slide and coverslip (x400) of lymph node aspirate (LNA) in cases where cervical lymphadenopathy was present. A serosuspect who was positive on at least one parasitological examination was confirmed as a gHAT case, denoted by T.

**Fig. 2.**
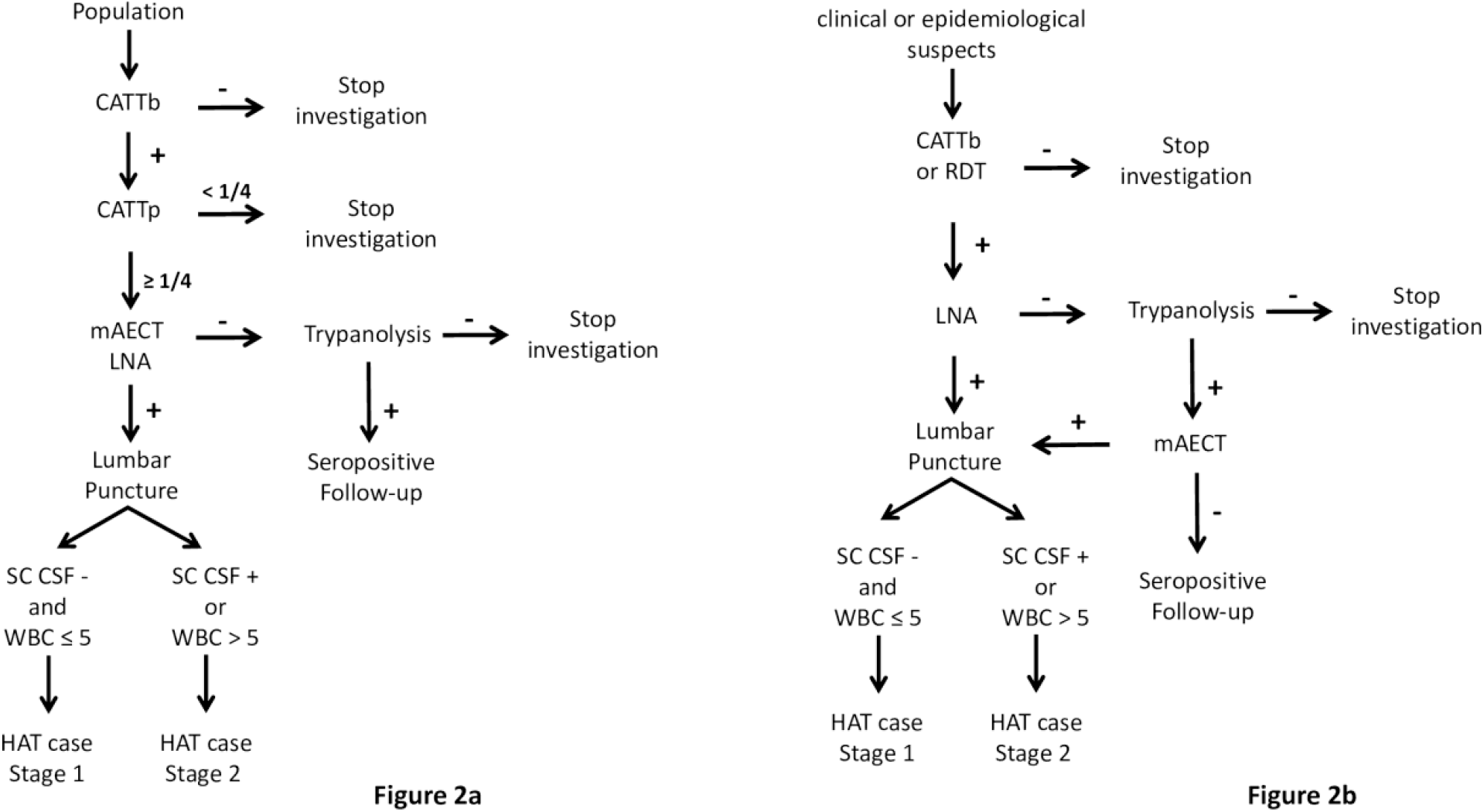
gHAT diagnosis algorithm for active screening and targeted active screening 2015-2019. Fig 2a. gHAT diagnosis algorithm for active screening Fig 2b. gHAT diagnosis algorithm for targeted active screening conducted during the “identification of villages at risk” strategy CATTb = card agglutination test for trypanosomiasis performed on whole blood; CATTp = card agglutination test for trypanosomiasis performed on plasma dilution; RDT = rapid diagnostic test; mAECT BC = miniature anion-exchange centrifugation technique performed using buffy coat; LNA = lymph node aspirate; SC-CSF = simple centrifugation of cerebrospinal fluid; WBC = CSF white blood cell /µl; -= negative; + = positive

Stage diagnosis was performed for all confirmed cases and was based on the technique of simple centrifugation of cerebrospinal fluid (SC-CSF) to allow visualisation of trypanosomes [17] and in the white blood cell (WBC) count in CSF (as WBC/µl). Cases who were negative for trypanosomes in CSF and with ≤ 5 WBC/µl were classified as having stage 1 disease, typically with mild or no symptoms. Cases who had trypanosomes visualised in CSF or with > 5 WBC/µl were considered to be in stage 2, typically with neurological disorders. People in stage 1 disease were treated with pentamidine and those in stage 2 with nifurtimox-eflornithine combination therapy (NECT) [18]. Post-treatment follow-up including lumbar puncture was performed only in the case of clinical suspicion of relapse as recommended by WHO [3].

Following some more recent case detections, targeted active screening strategies were put in place. These included door-to-door [8] and spatial follow-up of cases [19] by which it is possible to screen the family and most-at-risk populations that share the same daily spaces as the gHAT cases and unconfirmed serosuspects. The diagnostic algorithm is shown in Fig 2a.

Active screening was targeted at villages previously identified as being at greatest risk in each area based on historical, epidemiological and geographical data as defined by the so-called “identification of villages at risk” (IVR) strategy [20]. This strategy was mainly applied in the HDs at risk. During IVR activities, clinical and epidemiological suspects were tested with a simplified algorithm (Fig 2b). Serology was based on CATTb or a rapid diagnostic test (RDT SD1, Abbott Diagnostics, South Korea) [21] and parasitology was based only on LNA if enlarged lymph nodes were present. A sample of blood dried on filter paper was then collected to perform a trypanolysis (TL) test in the laboratory [22]. When the TL was positive, the village concerned was automatically selected for exhaustive active screening and the TL-positive subject was tested using mAECT BC.

Serosuspects with CATTp≥1/4 or positive RDT but negative in parasitology were tested with TL. TL-positive subjects were considered to be “seropositive” (denoted by S) and potential carriers of trypanosomes. They were followed once a year according to the algorithm in Fig 2a until the serological tests were negative or until parasitological confirmation and treatment.

### Passive screening

Passive screening refers to diagnosis made in fixed health facilities and was based on clinical suspicion, with the following symptoms considered suggestive of gHAT disease: 1) long-term fever and no effect of antimalarial treatment, 2) headache for a long period (> 14 days), 3) presence of enlarged lymph nodes in the neck, 4) severe weight loss, 5) weakness, 6) severe pruritus, 7) amenorrhea, abortion(s), or sterility, 8) psychiatric problems (aggressiveness, apathy, mental confusion, unusual increasing hilarity), 9) sleep disturbances (nocturnal insomnia and excessive daytime sleep), 10) motor disorders (abnormal movements, tremor, difficulty walking), 11) speech disorders, 12) convulsion, 13) coma [23]. All subjects in whom at least one of these symptoms was observed were considered clinical suspects. The algorithm described in Fig 2a was applied at the Projet de Recherches Cliniques sur la Trypanosomiase (PRCT, Daloa) reference center for the diagnosis and treatment of gHAT and the only sentinel site for passive screening in 2015.

In August 2017, passive screening was set up in 10 health centres of the endemic HDs of Bouaflé (Bonon focus) and Sinfra as part of the DiTECT-HAT research [23]. The diagnostic algorithm that was used is presented in Fig 3a. Clinical suspects were tested with three RDTs (SD1, HAT Sero-*K*-Set (Coris BioConcept, Belgium), and rHAT Sero-Strip (Coris BioConcept, Belgium)). Subjects who tested positive with at least one RDT were tested by parasitology (mAECT BC and LNA). Staging was performed on confirmed parasitological cases. TL on blood dried on filter paper [24] was then carried out on RDT-positive but parasitology-negative subjects. TL-positive subjects were considered seropositive.

**Fig. 3.**
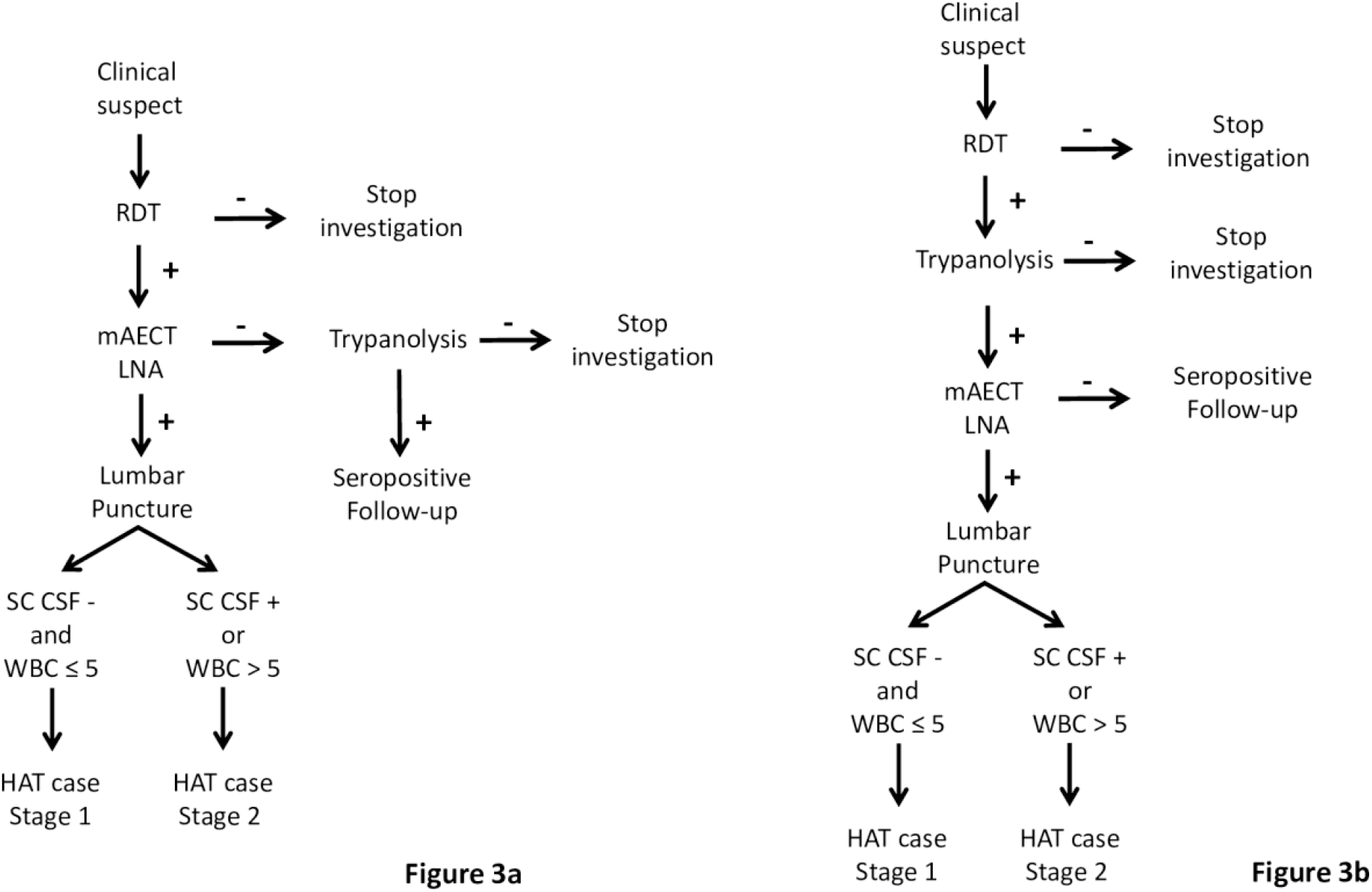
gHAT diagnosis algorithm for passive screening since 2015. Fig 3a. gHAT diagnostic algorithm for passive screening in endemic health districts Fig 3b. gHAT diagnostic algorithm for passive screening in hypo-endemic health districts and national coverage facilities RDT = Rapid Diagnostic Test; mAECT BC = miniature anion-exchange centrifugation technique performed with buffy coat; LNA = lymph node aspirate; SC-CSF = simple centrifugation of cerebrospinal fluid; WBC = CSF white blood cell /µl; -= negative; + = positive

In May 2018, passive screening was also set up in 13 health centres in the hypo-endemic HDs. The 13 health centres were selected based on epidemiological data (geographical distribution of the last cases detected) and their areas of influence, and in five neurology or psychiatry services in Bingerville (1), Bouaké (2) and Abidjan (2) (Fig 4) for national coverage. The diagnostic algorithm used was based on the RDT SD1 (Fig 3b). The 18 sites were supervised every three months and capacity building for doctors, nurses and laboratory technicians was carried out every year to optimise the effectiveness of the gHAT monitoring programme.

**Fig. 4.**
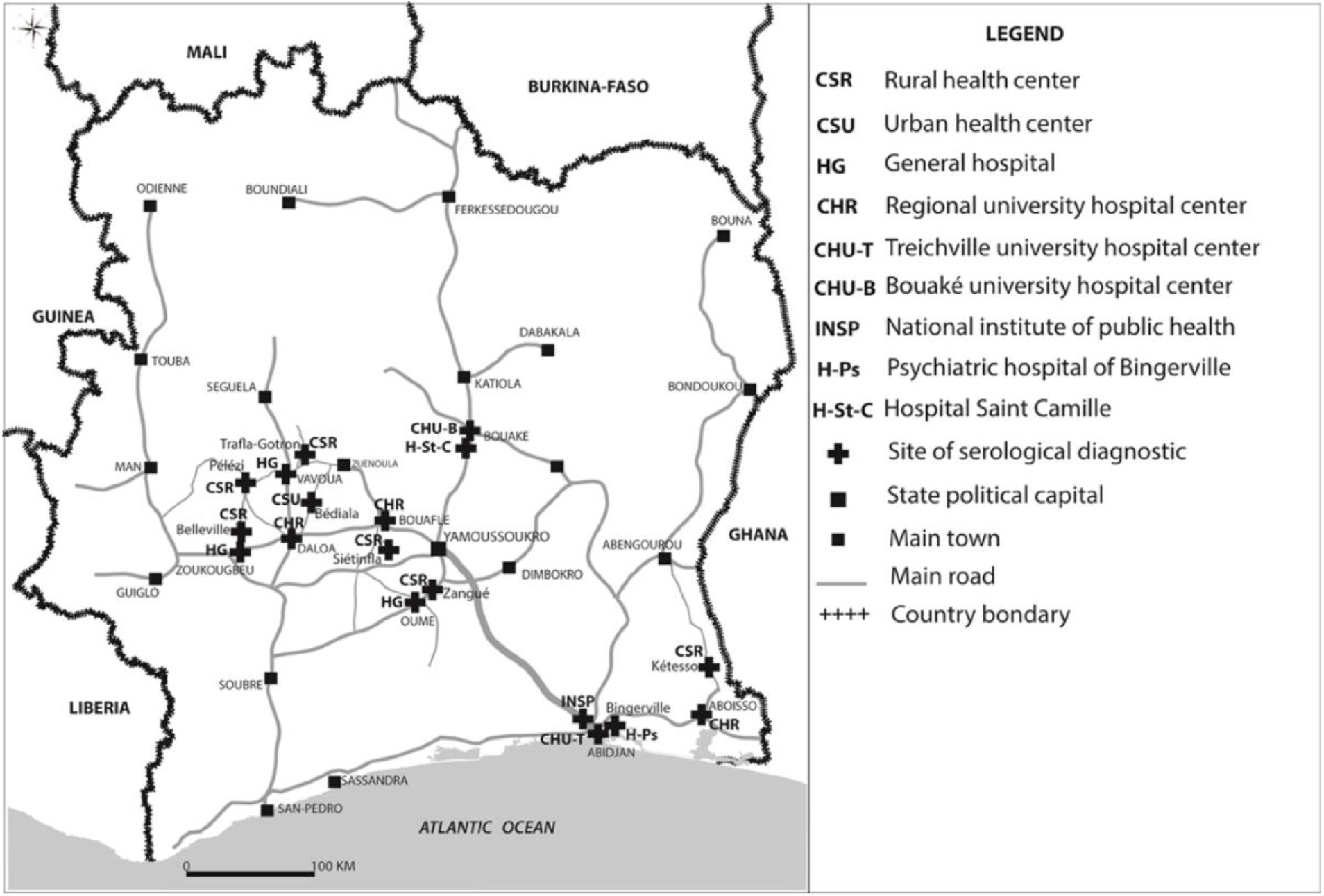
Sentinel sites for passive surveillance in hypo-endemic health districts and national coverage facilities 2015-2019. Source: Institut Pierre Richet, BNEDT/Openstreetmap.org

Strengthening of the capacities of the health workers, including the gHAT clinical suspicion and diagnosis, preceded the implementation of passive screening in these 28 gHAT sentinel sites.

### Vector control

Vector control (VC) provides a complementary method to screening and treatment and has been used to reduce tsetse populations and interrupt gHAT transmission in a variety of geographies (e.g. Chad, Democratic Republic of Congo, Guinea, and Uganda) [25,26,27]. In Côte d’Ivoire, VC mostly using Tiny Targets [28,29] but also Vavoua traps [30] both impregnated with deltamethrin, began in January 2016 in the HD of Bouaflé (Bonon focus). The first three years (until December 2018) of the Bonon intervention has been described elsewhere [31,32] but we summarise this and provide an update on the results obtained until December 2019. The first deployment in Bonon took place in February 2016 with 1,890 Tiny Targets. During annual redeployments in February 2017, February 2018 and February 2019, additional Tiny Targets were added to reach a total of 2,016 deployed in February 2019 (Fig 5a). It is during this redeployment that 57 Vavoua traps were set to reinforce VC in areas where tsetse were still being caught during periodic entomological assessments.

**Fig. 5.**
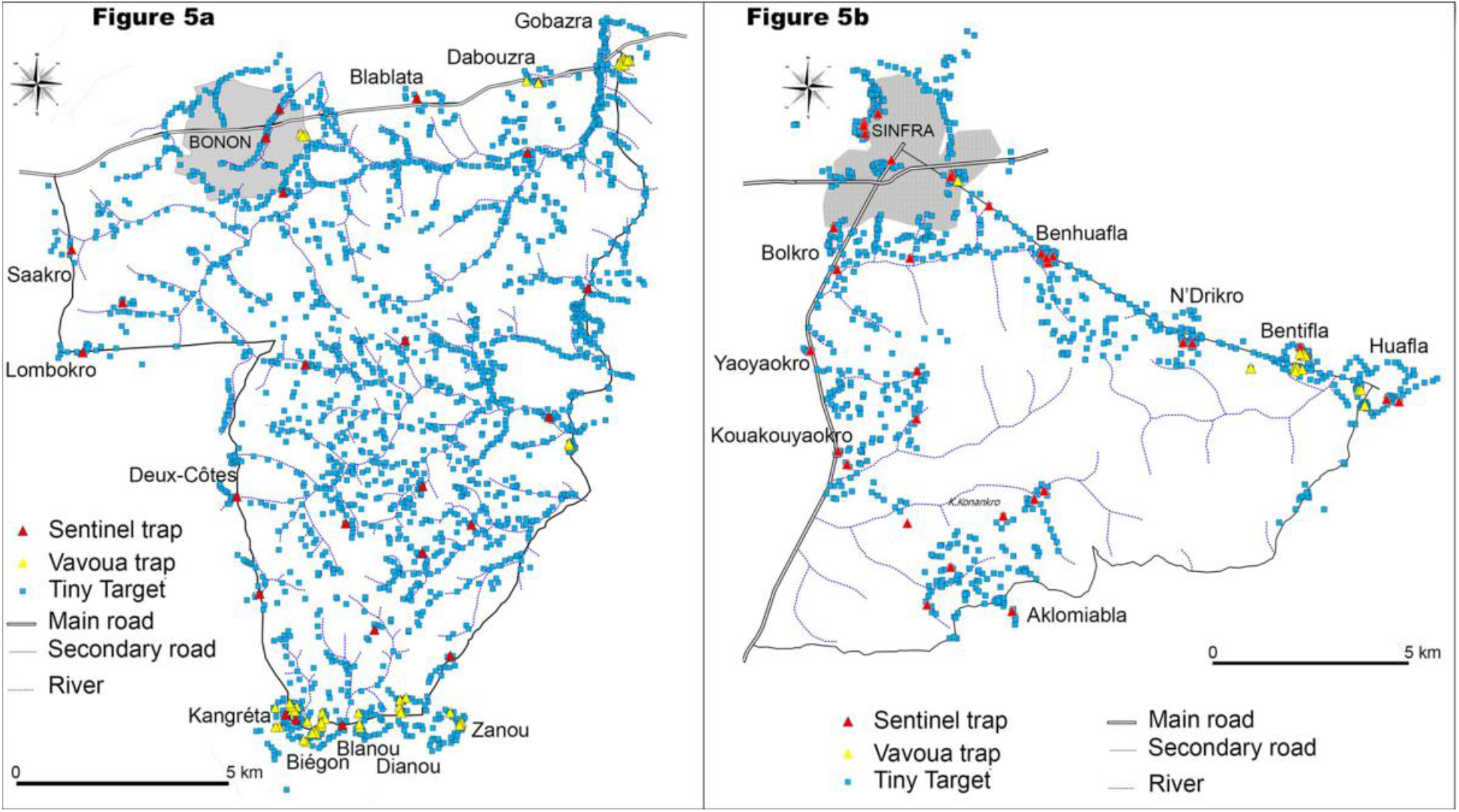
Distribution of control devices in endemic health districts. Fig 5a. Distribution of Tiny Targets and Vavoua traps in December 2019 and sentinel traps in the Bouaflé health district (Bonon focus) Fig 5b. Distribution of Tiny Targets and Vavoua traps in December 2019 and sentinel traps in the Sinfra health district Vavoua traps and Tiny Targets are impregnated control devices. Sentinel traps are unimpregnated Vavoua traps for capture. Source: Institut Pierre Richet, Openstreetmap.org

A more targeted VC began in May 2017 in the HD of Sinfra; the intervention aimed to control tsetse densities by deploying Tiny Targets supplemented by Vavoua traps at human/tsetse contact points in areas where the risk of transmission was believed to be highest. A total of 736 Tiny Targets were deployed in May 2017 and redeployed in July 2018 and July 2019. In July 2018, 115 additional Tiny Targets and 44 Vavoua traps were also deployed to reinforce VC in areas where tsetse were still being caught during periodic entomological assessments. In July 2019, the 44 Vavoua traps were replaced by Tiny Targets but 12 other additional Vavoua traps were set. At the end of 2019, 895 Tiny Targets and 12 Vavoua traps were deployed in the Sinfra HD (Fig 5b).

A T0 entomological survey using unimpregnated Vavoua traps for capture was conducted in the two foci before the first deployment of Tiny Targets. This T0 survey made it possible, in addition to sensitising local communities, (i) to delineate the intervention areas, (ii) to characterise the tsetse populations (species, densities, distribution) and (iii) to determine the distribution of control devices to be deployed. All traps were set for 48 or 96 hours and georeferenced using a GPS (Garmin 64®). Fly collection was made for two or four consecutive days and captured tsetse were identified by species with a magnifying glass and an identification key [33]. Apparent densities of flies per trap per day (ADTs) were determined.

To monitor the results of the VC campaign, sentinel traps (unimpregnated Vavoua traps for capture) were selected from those set during the T0 survey, on the basis of the ADT and to ensure homogeneous spatial coverage of the study area. These sentinel traps represented 10% of the total traps in T0. Quarterly entomological assessments used 30 sentinel traps in Bonon and 35 sentinel traps in Sinfra. All traps were set for 48 hours. Fly collection was made for two consecutive days and captured tsetse were identified by species [33]. ADTs were compared with those of T0.

### Modelling

For the modelling approach, we report on data for the period 2000–2021. S1 Data provides detailed active and passive case data partitioned by stage for the locations where there are sufficient data to conduct model fitting (Bonon, Bouaflé, Daloa, Oumé and Sinfra); only HDs with 10 or more data points (one data point is counted as a year with at least one passive case detected, or one active screening) were included as fewer data points were not deemed sufficient to provide a robust model fit. Other HDs are not included in that file and are omitted from model fitting, but aggregate case data (totals per year per location) are included in S1 Table. Following implementation of the different interventions and data collection we used mathematical modelling to assess these data and provide quantitative analysis of progress in Côte d’Ivoire. The modelling was used to estimate the underlying number of new infections each year between 2000 and 2021 and therefore assess the reduction in transmission over this time period. To achieve this, a previously developed mechanistic model (the “Warwick gHAT model”), originally published as [34], and more recently updated for the DRC [35] and Chad [36] was adapted for the context in Côte d’Ivoire and fitted to the annual time series data collected by PNETHA. The model has recently been described in detail elsewhere [35], and model equations and parameters are given in the S1 Text. Briefly, this model captures the natural history of infection in humans from exposure to the parasites and the relatively long progression through stage 1 and, if not treated, stage 2 infection. We simulate detection and treatment of cases via both active and passive screening, with the number of active cases identified within a year linked to the number of people tested by mobile teams. Tsetse are explicitly included in the model to capture human-tsetse-human transmission cycles and, furthermore, it is assumed that there is differential risk in exposure to tsetse bites between different people. As described in the vector control section, from 2016 and 2017, vector control was implemented in Bonon (Bouaflé HD) and Sinfra, respectively, and the corresponding reduction in tsetse populations is included in the transmission model for these foci. Tsetse trap data were used to inform the parameters associated with observed tsetse population reduction by fitting the tsetse population dynamic sub-model to these data via maximum likelihood estimation (see S1 Text).

To fit the full deterministic epidemiological model to data we use a Markov chain Monte Carlo (MCMC) methodology (see S1 Text) which compares annual active and passive case reporting simulated in the model to those observed in the data for each year for each HD. We carried out fitting for the two endemic HDs – Bouaflé and Sinfra – and two hypo-endemic HDs –Daloa and Oumé. Other listed hypo-endemic HDs and the “at-risk” HDs were not included in model fitting due to insufficient data points. As Bouaflé HD is comprised of two epidemiological foci – Bouaflé and Bonon – and only one of these had the vector control intervention deployed there, for the Bouaflé HD fitting we considered these small geographical units independently before aggregating to HD level.

The fitted model parameters are found in S1 Text Table D and include the basic reproduction number (*R0*), the relative risk of high-risk people being bitten by tsetse compared to low-risk people (*r*), the proportion of the population who are low risk (*k1*), the proportion of stage 2 cases that go on to be reported (as opposed to those that die undetected) (*u*), and passive detection rates (*ηH*, ***γ****H*). In some years the number of people tested in active screening was unknown, consequently this value was inferred during fitting. All these parameters were fitted independently in each region as they are assumed to be geographically variable. Prior parameter distributions for these fitted parameters and fixed parameter values can be found in the S1 Text Tables D and C, respectively.

In addition to inferring model parameters, the fitting process enabled us to estimate the expected number of annual new infections in each HD during the data period (2000–2021) and therefore to quantify the reduction in transmission over time. It also allowed us to calculate the probability that each location had achieved local EoT by a given year.

### Ethics

The HAT elimination project in Côte d’Ivoire (ElimTrypCI 2016–2019) received ethical clearance from the Comité National d’Ethique de la Recherche, Ministry of Public Health and Hygiene in Côte d’Ivoire (reference 030-18/MSHP/CNER-kp).

## Results

### gHAT situation during the 2000–2014 period

With the results of this article focusing on the 2015–2019 period, it is apt to describe the epidemiological situation observed before this period, based on the number of cases reported between 2000 and 2014 by the PNETHA (S1 Table). A total of 650 gHAT cases were reported, most of them from the Bonon subprefecture (323) and Sinfra HD (176) where the last two epidemics were recorded: early 2000s for Bonon [37,38] and mid-1990s for Sinfra [39,40]. During this period, 151 cases were recorded in the hypo-endemic HD (from one case in Gagnoa, Issia and Zuénoula HD to 50 cases in the Daloa HD). In all HDs, the number of cases gradually decreased over time. The last cases were recorded before 2006 in Aboisso, Gagnoa, Issia and Zuénoula HD, in 2008 in Zoukougbeu, in 2012 in Vavoua and in 2013 in Bouaflé subprefecture, while some cases were still reported in 2014 in the Bonon subprefecture, Sinfra, Daloa and Oumé.

### Active screening during 2015–2019

The results of exhaustive active screening activities carried out between 2015 and 2019 in the endemic HDs of Bouaflé and Sinfra and the hypo-endemic HD of Aboisso are presented in Table 1. A total of 13,074 people were tested, a single confirmed case of gHAT was detected in the HD of Bouaflé in 2015 and four seropositives were identified (three in Bouaflé and one in Sinfra). The results of the exhaustive active screening activities carried out between 2017 and 2019 in the HDs at risk are presented in S2 Table. A total of 28,796 people were tested and no gHAT cases or seropositive individuals were identified. The results of targeted active screening activities using the IVR method by HD between 2017 and 2019 are presented in S3 Table. A total of 5,093 clinical and epidemiological suspects were tested but no cases or seropositives were identified.

**Table 1.** Results of active screening activities in endemic and hypo-endemic health districts.

| **Health district (year)** | **PE<sub>x</sub>** | **S** | **T** |
| --- | --- | --- | --- |
| Bouaflé (2015) | 5744 | 3 | 1 |
| Sinfra (2016) | 3415 | 1 | 0 |
| Aboisso (2018) | 3915 | 0 | 0 |
| **Total** | **13074** | **4** | **1** |

PEx = population examined; S = number of seropositive subjects; T = number of confirmed gHAT cases

Follow-up results of seropositive subjects are shown in Table 2. A total of 97 subjects were followed and tested. One case was detected in 2017 in the district of Bouaflé and one in 2019 in Sinfra. In 2019, 18 subjects were still serologically positive and four of them were positive for both CATTp and TL.

**Table 2.** Results of follow-up of seropositive subjects.

| **Year** | **2015** |  |  | **2017** |  |  | **2108** |  |  | **2019** |  |  |
| --- | --- | --- | --- | --- | --- | --- | --- | --- | --- | --- | --- | --- |
| **Health district** | **PEx** | **S** | **T** | **PEx** | **S** | **T** | **PEx** | **S** | **T** | **PEx** | **S** | **T** |
| Bouaflé | 13 | 10 | 0 | 18 | 13 | 1 | 16 | 9 | 0 | 17 | 9 | 0 |
| Sinfra | 0 | 0 | 0 | 9 | 7 | 0 | 2 | 1 | 0 | 13 | 8 | 1 |
| Vavoua | 0 | 0 | 0 | 0 | 0 | 0 | 0 | 0 | 0 | 1 | 0 | 0 |
| Daloa | 2 | 2 | 0 | 2 | 2 | 0 | 2 | 1 | 0 | 2 | 1 | 0 |
| **Total** | **15** | **12** | **0** | **29** | **22** | **1** | **20** | **11** | **0** | **33** | **18** | **1** |

PEx = population examined; S = number of seropositive subjects; T = number of confirmed gHAT cases

Table 3 presents the results of active screening activities targeted at populations sharing the same spaces as the last gHAT cases and seropositives identified mainly in the HDs of Sinfra and Bouaflé, considering the results of spatial follow-ups in particular. A total of 3,105 people were tested between 2017 and 2019 but no cases or seropositive individuals were identified.

**Table 3.** Results of targeted active screening.

| **Year** | **2017** |  |  | **2018** |  |  | **2019** |  |  |
| --- | --- | --- | --- | --- | --- | --- | --- | --- | --- |
| **Health District** | **PEx** | **S** | **T** | **PEx** | **S** | **T** | **PEx** | **S** | **T** |
| Bouaflé | 121 | 0 | 0 | 204 | 0 | 0 | 1327 | 0 | 0 |
| Sinfra | 558 | 0 | 0 | 112 | 0 | 0 | 678 | 0 | 0 |
| Vavoua | 25 | 0 | 0 | 0 | 0 | 0 | 3 | 0 | 0 |
| Daloa | 64 | 0 | 0 | 0 | 0 | 0 | 13 | 0 | 0 |
| **Total** | **768** | **0** | **0** | **316** | **0** | **0** | **2021** | **0** | **0** |

PEx = population examined; S = number of seropositive subjects; T = number of confirmed gHAT cases

### Passive screening during 2015–2019

Between 2015 and 2019 a total of 169 people were tested and two cases of gHAT detected at the PRCT of Daloa, the reference centre for the diagnosis and treatment of gHAT recognized as such for decades by the populations of the HAT foci of the West Central Côte d’Ivoire [9]. Both of the cases were detected in 2015 from the 33 people tested that year. One of the two cases was from the HD of Sinfra and the other from the HD of Bouaflé. No further cases were detected from the 136 people tested at PRCT Daloa between 2016 and 2019.

Table 4 presents the results of the passive screening implemented between 2017 and 2019 in the endemic HDs of Sinfra and Bouaflé. A total of 3,433 clinical suspects were tested and two cases were reported in 2017, one in Sinfra and one in Bouaflé. They were diagnosed as stage 2 infections (the case in Sinfra was very advanced) as described in Koné et al. (2021). A third person, positive with the three RDTs and TL identified in Bouaflé in 2018, died following a sudden neurological deterioration without it being possible to confirm the gHAT diagnosis using parasitology. This case was considered a serological gHAT case in the PNETHA registers. No cases or seropositives were identified in 2019.

**Table 4.**
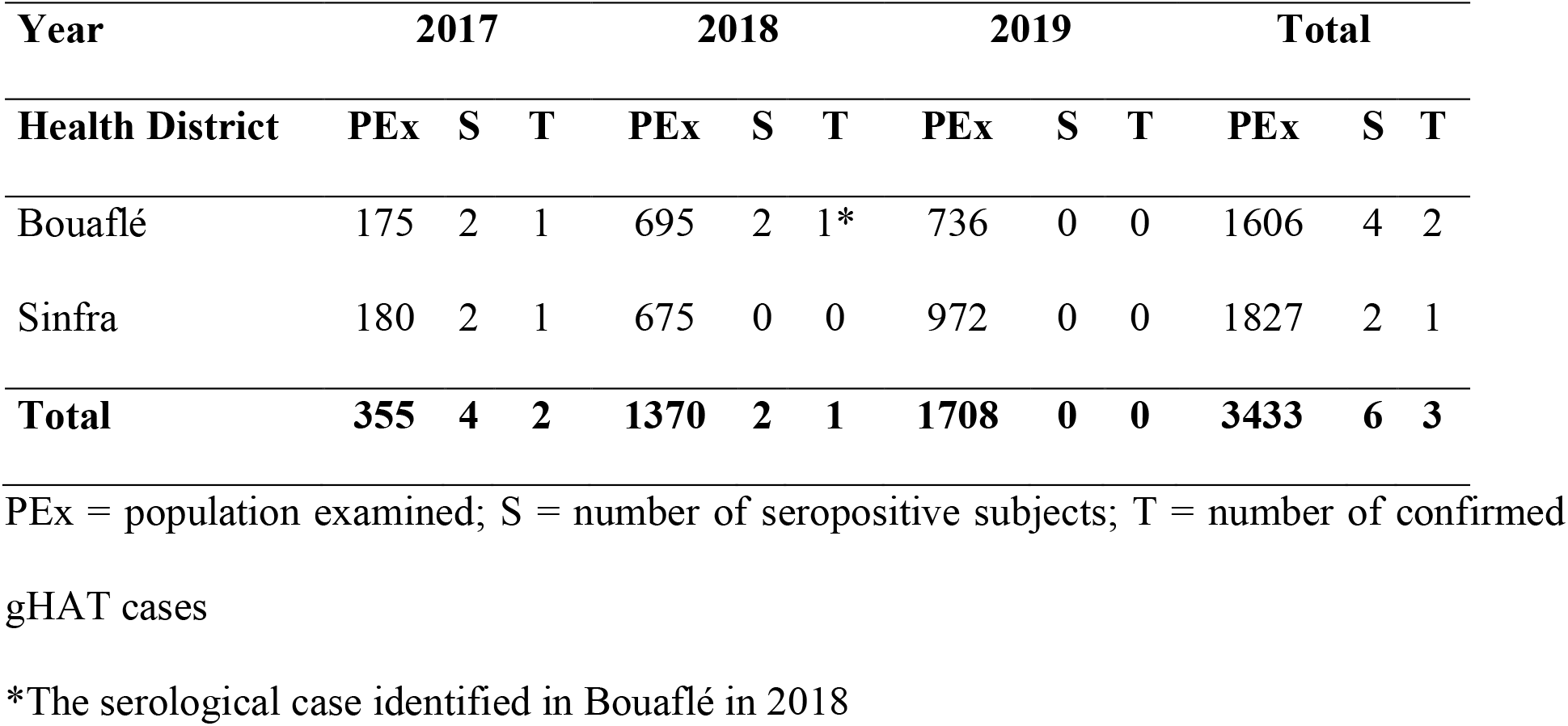
Results of passive screening in the endemic health districts.

| **Year** | **2017** |  |  | **2018** |  |  | **2019** |  |  | **Total** |  |  |
| --- | --- | --- | --- | --- | --- | --- | --- | --- | --- | --- | --- | --- |
| **Health District** | **PEx** | **S** | **T** | **PEx** | **S** | **T** | **PEx** | **S** | **T** | **PEx** | **S** | **T** |
| Bouaflé | 175 | 2 | 1 | 695 | 2 | 1* | 736 | 0 | 0 | 1606 | 4 | 2 |
| Sinfra | 180 | 2 | 1 | 675 | 0 | 0 | 972 | 0 | 0 | 1827 | 2 | 1 |
| **Total** | **355** | **4** | **2** | **1370** | **2** | **1** | **1708** | **0** | **0** | **3433** | **6** | **3** |

PEx = population examined; S = number of seropositive subjects; T = number of confirmed gHAT cases
\*The serological case identified in Bouaflé in 2018

The results of the passive screening implemented in 2018 and 2019 as part of sentinel site surveillance are shown in S4 Table. A total of 605 clinical suspects were tested, including 84 in national coverage facilities and 521 in hypo-endemic HDs. While five individuals were RDT-positive in hypo-endemic HDs, no cases or seropositives were identified.

A case of gHAT was detected and treated in 2018 in Koudougou in Burkina Faso as part of the passive surveillance set up there. The epidemiological investigation showed that this case was most likely infected in the outbreak of Bonon. The clinical questionnaire revealed significant neurological damage linked to an infection dating back several years. The case was included in the PNETHA registers as a confirmed case in 2018 from the HD of Bouaflé.

Therefore, in total, nine cases of gHAT were detected between 2015 and 2019 that were likely to have been infected in Côte d’Ivoire, six in the HD of Bouaflé and three in that of Sinfra (Table 5). The distribution of these cases aggregated by the method of screening and the stage of the disease is given in Tables 6 and 7. Of these nine cases, three were found in active screening, including one in exhaustive active screening, and two during the follow-up of seropositives. The six other cases were detected passively, including two at the PRCT in Daloa, three during surveillance in the HDs of Sinfra and Bouaflé and one during passive surveillance in Koudougou in Burkina Faso. The nine cases were diagnosed as stage 2 infections. Those detected in Côte d’Ivoire were treated with NECT. The case detected in Koudougou was treated with α-difluoromethylornithine (DFMO) according to the national procedure in Burkina Faso.

**Table 5.** Number of gHAT cases detected per year between 2015 and 2019.

| Number of gHAT cases / year |  |  |  |  |  |  |
| --- | --- | --- | --- | --- | --- | --- |
| Health District | 2015 | 2016 | 2017 | 2018 | 2019 | Total |
| Bouaflé | 2 | 0 | 2 | 2* | 0 | 6 |
| Sinfra | 1 | 0 | 1 | 0 | 1 | 3 |
| **Total** | **3** | **0** | **3** | **2** | **1** | **9** |

\* including the gHAT case diagnosed in Burkina Faso

**Table 6.** Method of screening of the gHAT cases detected between 2015 and 2019.

| Number of gHAT cases |  |  |  |  |  |  |  |  |  |  |
| --- | --- | --- | --- | --- | --- | --- | --- | --- | --- | --- |
| Year | 2015 |  | 2016 |  | 2017 |  | 2018 |  | 2019 |  |
| Health District | A | P | A | P | A | P | A | P | A | P |
| Bouaflé | 1 | 1 | 0 | 0 | 1 | 1 | 0 | 2* | 0 | 0 |
| Sinfra | 0 | 1 | 0 | 0 | 0 | 1 | 0 | 0 | 1 | 0 |

\* including the gHAT case diagnosed in Burkina Faso
A = Active detection; P = Passive detection

**Table 7.** Stage of the disease of the gHAT cases detected between 2015 and 2019.

| Number of gHAT cases |  |  |  |  |  |  |  |  |  |  |
| --- | --- | --- | --- | --- | --- | --- | --- | --- | --- | --- |
| Year | 2015 |  | 2016 |  | 2017 |  | 2018 |  | 2019 |  |
| Health District | S1 | S2 | S1 | S2 | S1 | S2 | S1 | S2 | S1 | S2 |
| Bouaflé | 0 | 2 | 0 | 0 | 0 | 2 | 0 | 2* | 0 | 0 |
| Sinfra | 0 | 1 | 0 | 0 | 0 | 1 | 0 | 0 | 0 | 1 |

\* including the gHAT case diagnosed in Burkina Faso; S1 = Stage 1; S2 = Stage 2

### Data according to the national indicator

Table 8 gives data for the national indicator for EPHP as defined by the WHO (average number of gHAT cases per year over 5 consecutive years/10,000 inhabitants, by HD, Franco et al., 2022). Only the HDs of Bouaflé and Sinfra are shown as these are the only HDs in which cases were reported between 2015 and 2019. Relative to the total population of the two HDs, the indicator was far below 1/10,000, a necessary condition for validating the EPHP.

**Table 8.** Number of gHAT cases according to the national indicator as defined by the WHO.

| Health District | gHAT cases | gHAT cases | gHAT cases | gHAT cases | gHAT cases | Average cases/year | Average population* | n/10,000 inhabitants |
| --- | --- | --- | --- | --- | --- | --- | --- | --- |
|  | 2015 | 2016 | 2017 | 2018 | 2019 |  |  |  |
| Bouaflé | 2 | 0 | 2 | 2 | 0 | 1,2 | 725,778 | 0,017 |
| Sinfra | 1 | 0 | 1 | 0 | 1 | 0,6 | 354,954 | 0,017 |

\* Source: Institut National de Statistique, Recensement Général de la Population et de l'Habitat (RGPH) 1998 and 2014

### Vector control

In Bonon, 267 traps were set during the T0 survey carried out in June 2015. In total, 1,894 flies of the species *Glossina palpalis palpalis* were captured, *i*.*e*. an average ADT of 3.54 flies/trap/day. No other tsetse species was caught. The average ADT of the 30 sentinel traps (1,322 tsetse captured) was 22.03 flies/trap/day. It was 0.25 flies/trap/day (15 tsetse captured) during the 15^th^ entomological evaluation (T15, December 2019), *i*.*e*. a reduction of 98.9% in tsetse density (Fig 6a). In Sinfra, the T0 survey was carried out in November 2016 with 339 traps set and 988 flies of the species *G. p. palpalis* captured (ADT 1.45 flies/trap/day). No other tsetse species was caught. The average ADT of the 35 sentinel traps was 8.99 flies/trap/day with 866 tsetse captured. It was 0.11 flies/trap/day (8 tsetse captured) during the 10^th^ entomological evaluation (T10, December 2019), *i*.*e*. a reduction of 98.8% in tsetse density (Fig 6b).

**Fig. 6.**
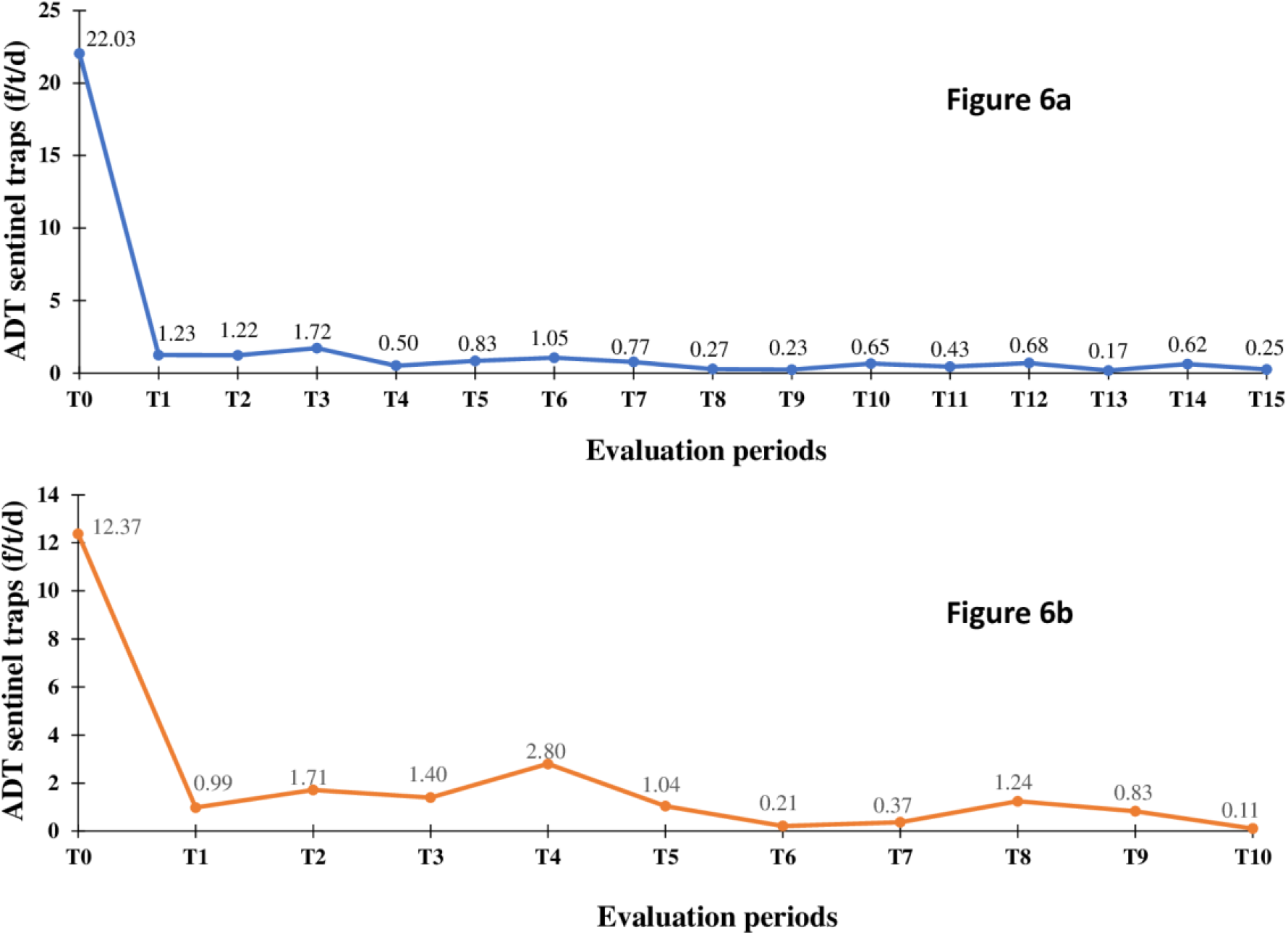
Evolution of the tsetse apparent density per trap during the entomological evaluations. Fig 6a. Evolution of the tsetse apparent density per trap during the entomological evaluations from T0 (June 2015) to T15 (December 2019) in the Bouaflé health district (Bonon focus) Fig 6b. Evolution of the tsetse apparent density per trap during the entomological evaluations from T0 (November 2016) to T10 (December 2019) in the Sinfra health district ADT = Apparent Density per Trap; f/t/d = tsetse flies/trap/day

### Modelling

Fig 7 shows the aggregated results for fitting the deterministic model to the longitudinal gHAT case data for Bouaflé, Sinfra, Oumé and Daloa HDs for the 2000–2021 period. Individual HD fits (or subprefecture fits in the case of Bouaflé HD) can be found in the S1 Text. Much of the case reporting dynamics appear to be driven by Bonon subprefecture of Bouaflé HD which accounted for between 19 and 100% of all actively reported cases and 11–62% of passively reported cases during 2000–2008 for the fitted foci. Similarly, Bonon subprefecture was estimated to contribute 25–56% of annual new infections. Passive case reporting in Sinfra occurred at a similar level to Bonon (24–67% of cases between 2000–2008) and saw a slow but steady decline. Active case reporting in Sinfra mirrored the pattern in active screening coverage during the 2000s. Daloa and Oumé HDs and Bouaflé subprefecture all had very low case reporting, never reaching more than 14 cases per year per foci across 2000–2021; correspondingly the model fitting produced very low estimates of annual new infections.

**Fig. 7.**
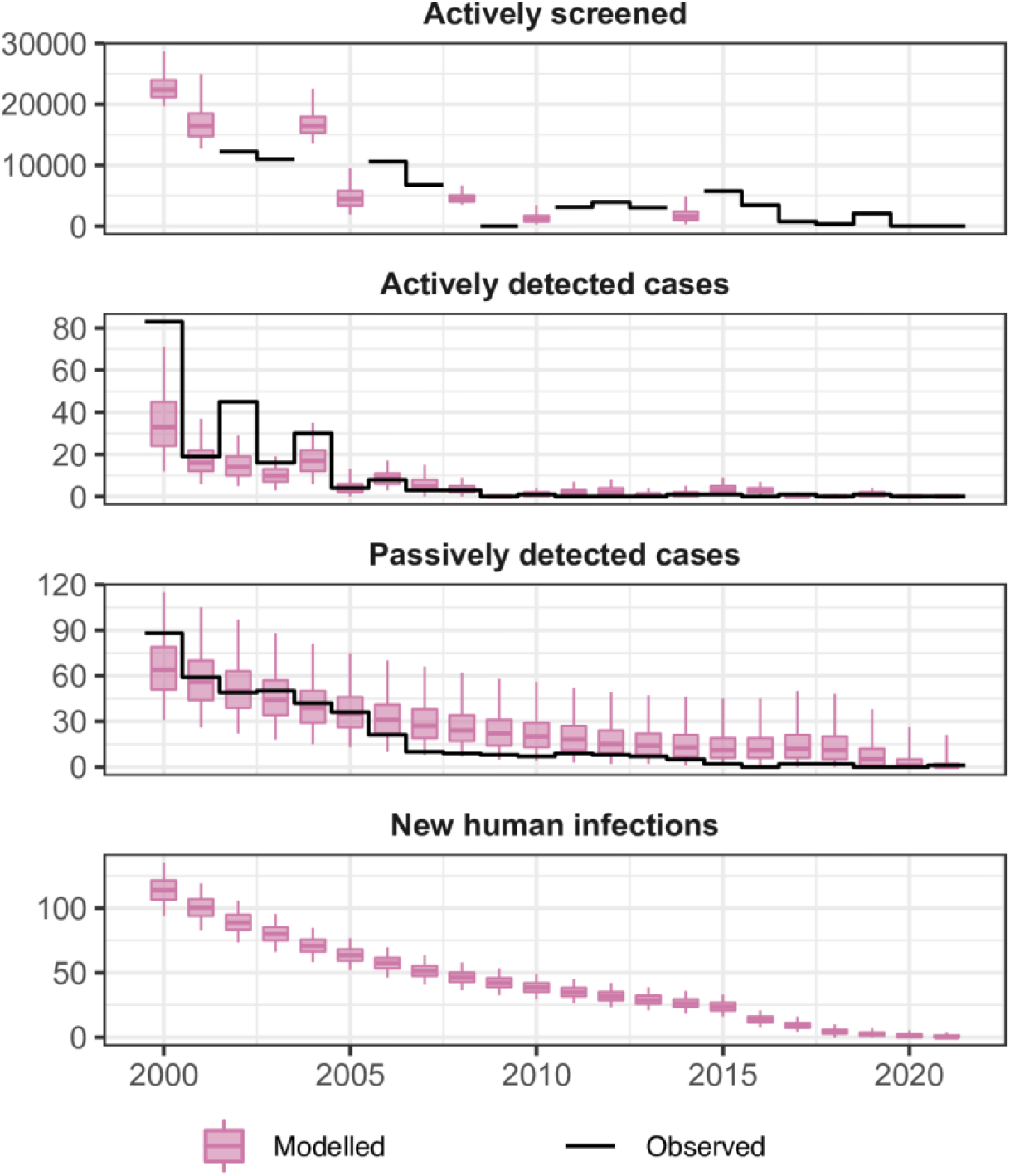
Aggregate fits to the data from Bouaflé, Sinfra, Oumé and Daloa health districts between 2000–2021. The black lines show the observed data (either number of people screened or cases) and the pink box and whisker plots show the deterministic model with stochastic sampling (centre line is the median, boxes contain 50% credible intervals (CIs) and whiskers show 95% CIs). Some years of data are missing the total number of people actively screened so this was estimated during fitting with results shown as box and whiskers. New infections are not directly observable and are estimated through the model based on case reporting.

By 2011 there was extremely little case reporting across all foci. Fig 8 shows the year in which transmission was estimated to be interrupted for each HD – for this calculation we utilised the analogous stochastic version of our model and the posterior parameterisation to better factor in chance events around EoT and remove the need to use a proxy threshold to compute EoT using deterministic outputs (see S1 Text for more details). In the Bouaflé subprefecture, Daloa, and Oumé, we computed that there was a moderate probability of EoT occurring in 2015 or earlier, however there is some considerable uncertainty in the year of EoT in these locations. In Bonon and Sinfra the use of highly impactful VC (from 2016 and 2017, respectively) coupled with the low or zero case reporting in recent years, results in model estimates of 2016 and 2018 for the average year of EoT (see Fig 8 and S1 Text Fig M) and these estimates have less uncertainty than the other HDs. Our modelling estimated that 52–71% of the transmission reduction across HDs likely occurred between 2000–2010, however it was after 2010 that transmission was likely interrupted (See Table 9 and S1 Text Table H).

**Table 9.** Estimated transmission reduction for Bouaflé, Sinfra, Oumé and Daloa health districts.

| Health district | Transmission reduction |  |  |
| --- | --- | --- | --- |
|  | 2000–2010 | 2011–2021 | 2000–2021 |
| Bouaflé | 70.9% [60.0–79.6%] | 100% [100–100%] | 100% [100–100%] |
| Daloa | 54.49 [24.4–73.9%] | 100% [61.1–100%] | 100% [74.9–100%] |
| Oumé | 52.0% [20.6–73.5%] | 100% [62.8–100%] | 100% [75.4–100%] |
| Sinfra | 67.0% [47.8–80.0%] | 100% [100–100%] | 100% [100–100%] |

**Fig. 8.**
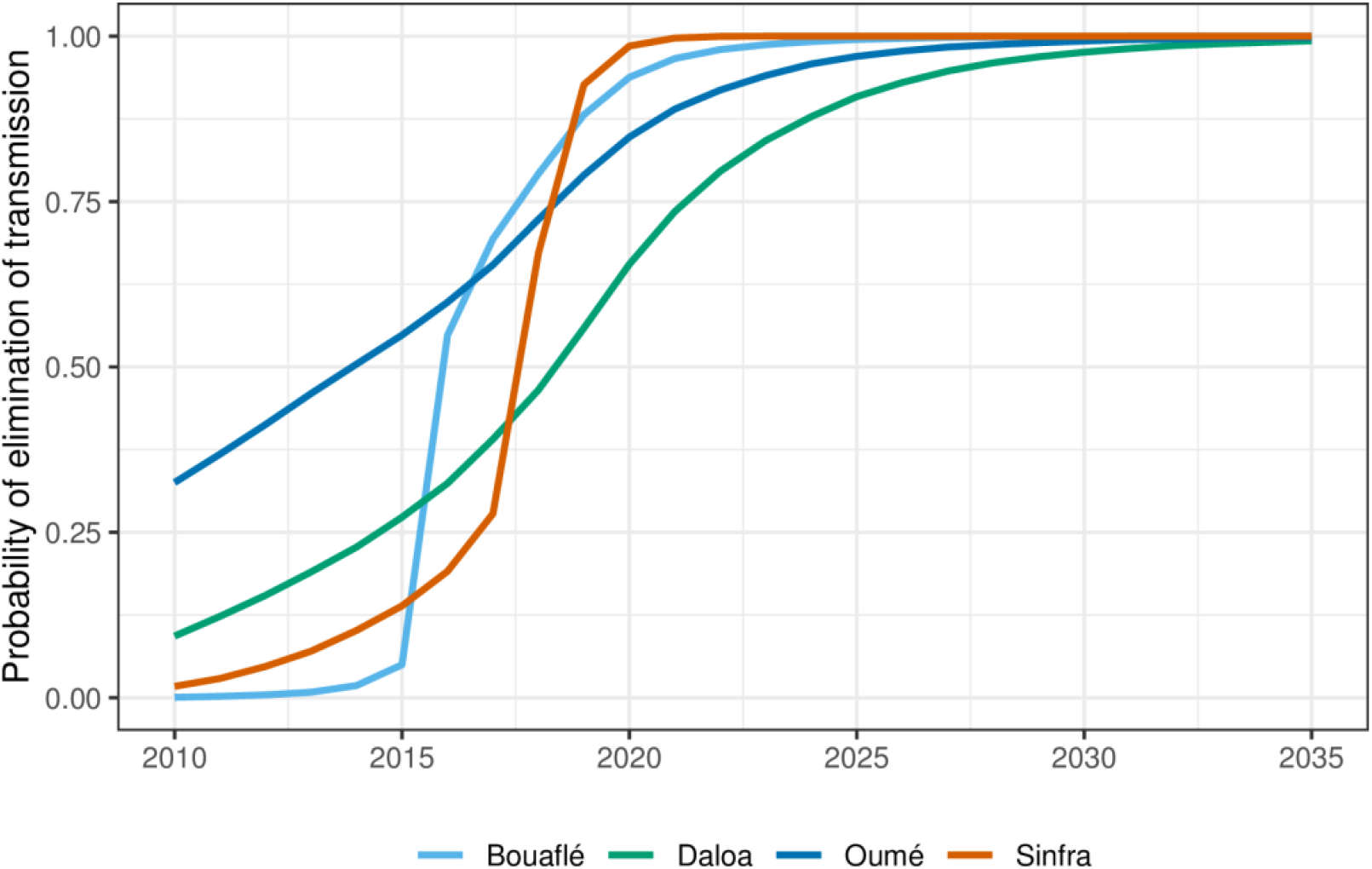
Estimated probability of elimination of transmission (EoT) by year for Bouaflé, Sinfra, Oumé and Daloa health districts. The probability is computed by assessing the proportion of stochastic model simulations where there are zero new transmission events for that year and the subsequent years for each year for each location. For disaggregation of Bouaflé HD into Bonon and Bouaflé subprefectures see S1 Text.

The reductions are computed through model fitting to historical data with the deterministic model. Medians and 95% credible intervals are given.

## Discussion

gHAT control activities in Côte d’Ivoire have been based on an integrated approach, consisting of a combination of medical interventions (active and passive screening followed by treatment) and vector control. The results of active screening and identification of villages at risk have shown that there is most likely very little or no transmission of *T. b. gambiense* in at-risk HDs. Indeed, no gHAT cases or seropositives were identified out of nearly 34,000 people tested between 2017 and 2019. Exhaustive and targeted active screening and passive screening activities also support the hypothesis of low or no transmission in hypo-endemic HDs with no cases detected even in the PRCT of Daloa.

The results of active screening have shown a clear reduction in the reported prevalence of the disease in the HDs of Bouaflé and Sinfra. They have also justified the gradual abandonment of exhaustive active screening in favour of targeted active screening and passive screening already described in several gHAT foci [41]. These strategies, however, confirmed that two HDs still had extant gHAT infections during 2015–2019. The results shown in the present study confirm the continued trend of a decrease in case reporting already observed since the beginning of the 2000s and the discovery of the last active focus of gHAT in Côte d’Ivoire [42,43], and this is despite the socio-political crisis that Côte d’Ivoire went through between 2002 and 2012 [9,38]. Monthly supervision and annual retraining of the health workers involved in this project have contributed greatly to the effectiveness of the implemented strategy.

Modelling suggests that there is a corresponding decrease in underlying transmission, and all HDs have a very high probability that EoT has already occurred in Côte d’Ivoire. Collected data confirm the importance of having adapted screening strategies by targeting areas and populations at risk and which made it possible to detect the majority of the remaining gHAT cases [8,23,44,45]. The fact that all the notified cases were in stage 2 of the disease indicates that these are likely to be relatively old infections and there is probably an absence of recent transmission.

The vector control carried out in the HDs of Bouaflé (Bonon focus) and Sinfra led to a sharp drop in tsetse densities from the first deployment of Tiny Targets and/or traps. A tsetse density reduction of more than 90% was rapidly achieved in each focus and maintained until the end of 2019. The presence of residual populations of tsetse was maintained in conserved forests consisting essentially of sacred forests (often on the outskirts of villages) in which the laying of screens and traps was often forbidden. These forests constitute favourable biotopes for tsetse, due to the presence of free ranging domestic pigs which frequent them regularly and constitute an ideal source of food [31,46,47], in addition to other possible hosts such as reptiles. Pigs have already been described as a preferential feeding host for *G. p. palpalis* [48,49], the only tsetse species present in the two vector control areas. Nevertheless, vector control is believed to have had a substantial impact on the risk of transmission, as has already been described for the Bonon focus [31] and is supported for both Bonon and Sinfra by the modelling analysis conducted as part of the present study.

The gHAT epidemiology in Côte d’Ivoire also depends on the gHAT situation in neighbouring countries. Côte d’Ivoire has a border with five endemic HAT countries: Liberia, Guinea, Mali, Burkina Faso and Ghana (Fig 1) with large cross-border mobilities that pose a risk of spreading gHAT from border countries to Côte d’Ivoire but also from Côte d’Ivoire to neighbouring countries. In the past, most of Côte d’Ivoire’s historic foci were in direct contact with foci in neighbouring countries [50]. But since 2000, no gHAT cases have been detected in a cross-border foci and no cases in Côte d’Ivoire appear to have been infected in a neighbouring country, although we cannot rule that out. Since 2015, very few cases have been reported from neighbouring countries in which there no longer seem to be active foci excepted on the Guinean coast [2], which is very far from Côte d’Ivoire. The risk of gHAT spread in Côte d’Ivoire from a neighbouring country is therefore very low. However, cases imported from Côte d’Ivoire have been regularly reported in Burkina Faso due to the large historical and recent population movements between these two countries [9,51,52]. However, the decrease in prevalence in Côte d’Ivoire has reduced the risk of spread to Burkina Faso and the case detected in Koudougou in 2018 (infected in the Bouaflé HD) is the latest reported.

It is important in this article to mention other phenomena that have not prevented the achievement of the EPHP of gHAT in Côte d’Ivoire but which should be considered as key in regard to the EoT. This is particularly the case of the role of a domestic or wild animal reservoir in the *T. b. gambiense* HAT epidemiology that is still under debate [53]. In Côte d’Ivoire, free-ranging pigs have been identified in the Sinfra, Bonon and Vavoua foci as a multi-reservoir of *T. brucei* and/or *T. congolense* with mixed infections of different strains [46,47]. This trypanosome diversity hinders the easy and direct detection of *T. b. gambiense*. It is important to stress both the lack of tools to prove or exclude with certainty the presence of *T. b. gambiense*, and the need of technical improvements to explore the role of pigs and animals in general, in the epidemiology of HAT.

A residual human reservoir of *T. b. gambiense* could also compromise the EoT in areas where tsetse are still occuring. Seropositive individuals (positive with either CATT or RDTs and with the highly specific trypanolysis test, but negative with parasitological tests) have been identified in both endemic HD (Bouaflé and Sinfra), and in some hypo-endemic HD. If we have already shown that some of them are spontaneous cure (and no longer pose a risk of transmission), we also observed that others are potential latent infections [22,54] and this is well illustrated by the two cases detected in 2017 and 2019 in Bouaflé and Sinfra HD, respectively. The case detected in Sinfra had been monitored for more than 20 years. Fortunately, he had been living in Abidjan for 15 years and probably did not pose any risk of transmission. In addition to these cases where infection is tolerated and diagnosis is difficult, there is also the difficulty of detecting gHAT cases in a context where the prevalence has become very low to the point that the disease is no longer considered a threat by the communities or by health workers. This is well illustrated by the complex health seeking pathway of the case passively diagnosed in 2017 in the Sinfra HD. in which the first disease symptoms appeared three years earlier and the patient had visited several health care centers and hospitals in different cities [45].

The modelling analysis presented here used a previously developed mechanistic model which explicitly incorporated human-tsetse contact and parasite transmission as well as heterogeneities in exposure of people to tsetse blood feeding. Longitudinal case data was used to parameterise the model for each geographical location and the resultant model fits align well with reported active and passive cases. Nevertheless, it is acknowledged that this model variant does not incorporate the possibility for non-human animal-tsetse transmission cycles, nor potentially self-curing asymptomatic human infections. Either of these two possibilities could lead to more transmission events per detected case, and therefore to more pessimistic model outcomes [55,56]. Despite this, the extremely low case reporting across several years in Côte d’Ivoire may indicate that these transmission routes (if they exist) are not sustaining transmission to humans; modelling analyses in the low-prevalence regions of the former Equateur province of the Democratic Republic of Congo [55] and the Mandoul focus of Chad [36] have found this kind of persistent low or zero reporting is suggestive of very limited or no infection contribution from non-human animals. Furthermore, in the foci with vector control, the large reduction in tsetse population density will have reduced transmission between tsetse and any potential infection source (animal or human).

The dynamic tsetse population sub-model used here includes the pupal stage of development as well as adult flies; this enabled us to model some resurgence of fly populations between Tiny Target deployments. This type of bounceback was included in the model to capture a plausible biological mechanisms for tsetse population growth between vector control deployment and this model matched fly catches well. We acknowledge that it is possible for bounceback to also occur through reinvasion of flies from neighbouring regions with no control and that other sources of tsetse-related data including habitat or climate data might be useful in trying to elucidate drivers of bounceback in different locations, especially after target deployments are stopped, or to predict potential pockets of high tsetse density, however these data require the use of alternative geostatistical modelling [57] which is beyond the scope of the present study.

While we use a stochastic simulation to model the human population, we have used a deterministic ODE-based approach to model tsetse dynamics. In general, a stochastic model would be preferred, especially at very low prevalence, however due to lack of data on the total tsetse population and inability to uncouple the size of tsetse population from the probability of infection per bite, we must instead fit a relative vector density [58]. This means that we are no longer modelling a discrete population of vectors, but a continuous density so a stochastic model is infeasible. Due to the slow dynamics of gHAT and short life-span of tsetse, however, we expect this to have minimal impact on our estimates of elimination. In this study the focus was on past transmission, however we do provide illustrative projections for the probability of EoT in Fig 8 and S1 Text Fig M. These projections assume the continuation of the current strategy in all health districts, however further work should be done to explore a range of plausible future strategies including scaling back. We recommend that these type of model projections are also coupled with health economic evaluations which could be used to assess what, how much and where investment is needed for the gHAT programme in Côte d’Ivoire to quantify pathway to country-wide EoT, verification of EoT, and also to consider what constitutes an efficient package of interventions to reach this target. As a preliminary study, a recent paper examined the costs of vector control using Tiny Targets in the Bonon focus from 2016 to 2017 [32].

This article summarises the information provided in the dossier that led to the WHO’s validation of EPHP in December 2020 [59]. This success was achieved through a one health approach combining medical surveillance and vector control interventions [12] and an integrated multi-stakeholder and multidisciplinary approach often needed in the fight against other infectious diseases including NTDs [60]. Research has played a major role in adapting tools and strategies to new epidemiological realities that present novel challenges. Moving towards the future, the strategies that will be put in place will have to be increasingly effective by targeting the areas and populations most at risk, to diagnose the last cases and minimise the risk of transmission via restriction of the human-tsetse and tsetse-*T. b. gambiense* contact.

The objective in Côte d’Ivoire is now to reach EoT by 2025. This requires continuing to adapt the control strategies. For the 2023-2025 step, focus will be on passive surveillance at the national scale and on reactive and targeted active screening including the follow-up of seropositive subjects and people who share their places of life. Medical and entomological capacities for reaction will be maintained, should any case be identified in the country. It is also crucial to consider some new challenges, including (i) the potential pig reservoir of *T. b. gambiense* and its consequences on HAT transmission, and (ii) community engagement to continue implementing suitable control strategies in a context where rare cases, if any, will be diagnosed. All the activities will be carried out in order to be able to compile the necessary information for the request for verification of EoT that will be submitted by the Ministry of Health to WHO in 2025.

## Data Availability

All aggregate case and screening data presented in the present work are contained in the manuscript. Code and results of the modelling analysis can be found online at https://osf.io/jtrs9/.

## Acknowledgements

The authors wish to thank their institutions, and all contributors to the control of gHAT in Côte d’Ivoire, whatever their status. The PNETHA and partners thank WHO for their support to collect and consolidate data on reported gHAT cases in the frame of the WHO Atlas. The authors wish to pay tribute to Pierre Cattand who significantly contributed to the HAT control in Côte d’Ivoire.

## Supporting information Captions

**S1 Table. Number of gHAT cases per Health District reported by the PNETHA during the 2000-2014 period** The Bouafle HD includes the Bonon HAT endemic foci. To be more precise, we present separately the data of the Bonon subprefecture (Bouaflé-Bonon subprefecture that includes the Bonon focus) and the rest of the Bouaflé HD (Bouaflé subprefecture). (xlsx)

**S2 Table. Results of the exhaustive active screening activities carried out between 2017 and 2019 in the Health District at risk** PEx = population examined; S = number of seropositive subjects; T = number of confirmed gHAT cases. (xlsx)

**S3 Table. Results of targeted active screening activities using the “identification of villages at risk” method by HD between 2017 and 2019** PEx = population examined; S = number of seropositive subjects; T = number of confirmed gHAT cases. (xlsx)

**S4 Table. Results of passive screening in the national coverage facilities and hypo-endemic health districts** PEx = population examined; RDT pos = number of positive rapid diagnostic tests; HD = Health District ; CHU = Centre Hospitalier Universitaire; INSP = Institut National de Santé Publique ; CHR = Centre Hospitalier Régional ; HG = Hôpital Général ; CSU = Centre de Santé Urbain ; CSR = Centre de Santé Rural. Locations for each are indicated in Fig 4. (xlsx)

**S1 Figure. Limits and status of HAT foci in Côte d’Ivoire until 2014** (pptx)

**S1 Text. Additional modelling methods and results** Details on the model definition, parameter inputs, fitting methodology, posterior parameter distributions following fitting, fits against the data. (pdf)

**S1 Data. Breakdown of gHAT cases per disease focus, screening mode and stage for the period 2000–2021 for locations where model fitting was performed** Data as used for the model fitting. This file does not include HDs with insufficient data for model fitting. (xlsx)

**S2 Data. Tsetse catch data and target deployment information for Bonon and Sinfra foci** Data used for fitting the tsetse population model in regions which implemented vector control. (xlsx)

