## Supplementary figures and images for "Towards the sustainable elimination of human African trypanosomiasis in Côte d’Ivoire using an integrated approach"

### S1 Figure

#### Slide 1
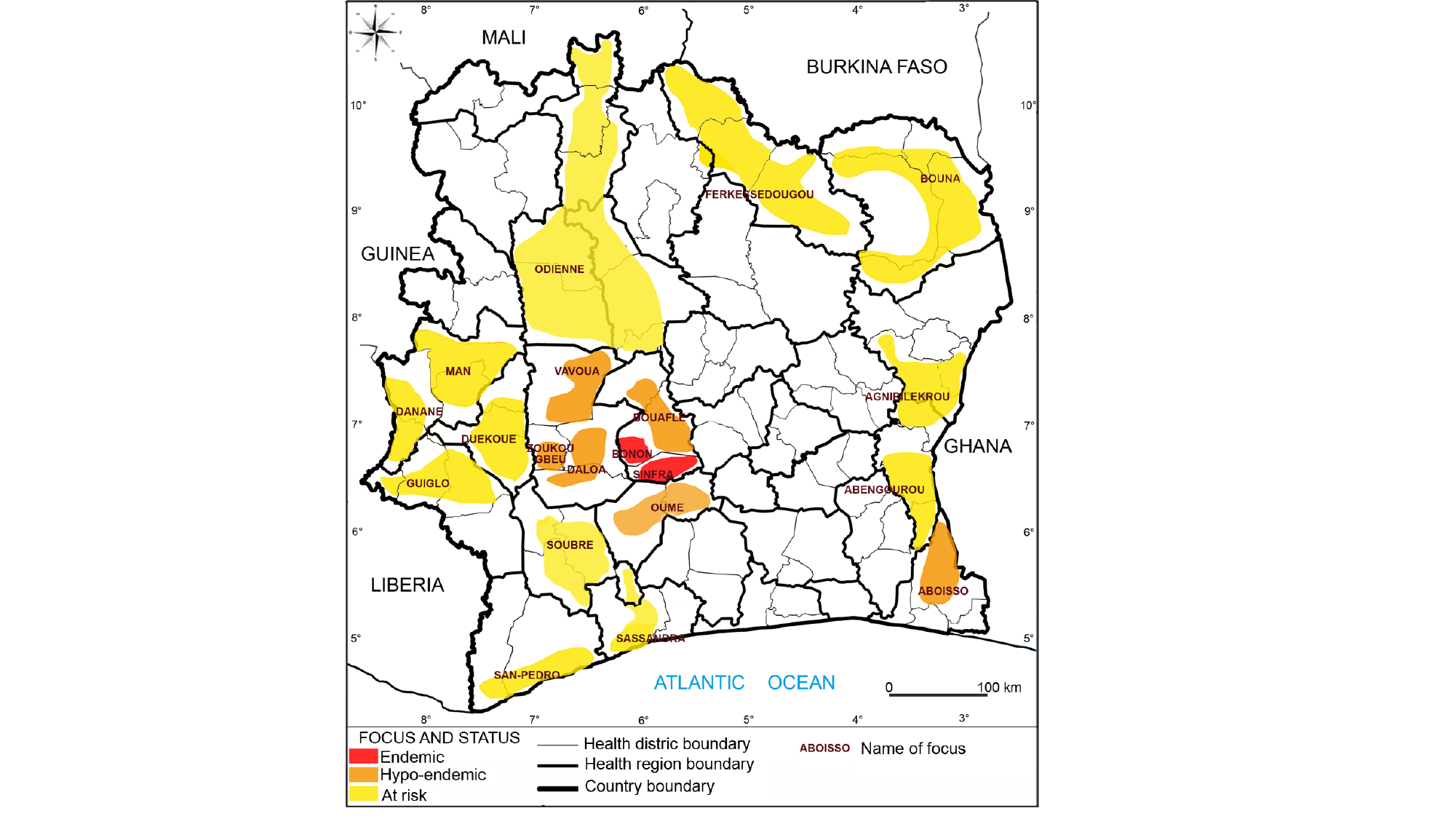
