## Supplementary material for "Towards the sustainable elimination of human African trypanosomiasis in Côte d’Ivoire using an integrated approach": S1 Text

### **Supplementary Information**: Towards the sustainable elimination of human African trypanosomiasis in Côte d'Ivoire using an integrated approach S1 Text: Additional modelling methods and results


Dramane Kaba[1], Mathurin Koffi[2], Lingué Kouakou[3], Emmanuel Kouassi N'Gouan[4], Vincent Djohan[1], Fabrice Courtin[5], Martial Kassi N'Djetchi[4], Bamoro Coulibaly[1], Guy Pacôme Adingra[1], Djakaridja Berté[1], Bi Tra Dieudonné Ta[1], Minayégninrin Koné[1], Samuel A Sutherland[6,7], Ron E Crump[6,8], Ching-I Huang[6,8], Jason Madan[6,7], Paul R Bessell[9], Antoine Barreaux[5], Philippe Solano[5], Emily H Crowley[6,8], Kat S Rock[6,8]◕, and Vincent Jamonneau[1,5]◕*

[1]Unité de Recherche « Trypanosomoses », Institut Pierre Richet, Bouaké, Côte d'Ivoire
[2]Laboratoire de Biodiversité et Gestion des Ecosystèmes Tropicaux, Unité de Recherche en Génétique et Epidémiologie Moléculaire, UFR Environnement, Université Jean Lorougnon Guédé, Daloa, Côte d'Ivoire
[3]Programme National d'Élimination de la Trypanosomiase Humaine Africaine, Abidjan, Côte d'Ivoire
[4]Projet de Recherches Cliniques sur la Trypanosomiase, Daloa, Côte d'Ivoire
[5]Unité Mixte de Recherche IRD-CIRAD 177, INTERTRYP, Institut de Recherche pour le Développement, Université de Montpellier, Montpellier, France
[6]Zeeman Institute for Systems Biology and Infectious Disease Epidemiology Research, Mathematical Sciences Building, The University of Warwick, Coventry, CV4 7AL, U.K.
[7]Warwick Medical School, The University of Warwick, Coventry, U.K.
[8]Mathematics Institute, Zeeman Building The University of Warwick, Coventry, CV4 7AL, U.K.
[9]Independent Consultant, Edinburgh, UK


January 21, 2023


◕These authors contributed equally to this work.


#### S1.1 Data

Population sizes for each health district or sub-prefecture used for fitting are given in Table A. Epidemiological data used to fit the model consist of the number of cases detected by active screening, the number of cases detected by passive screening and the population actively screened in each focus in each year from 2000–2021. Some earlier years have incomplete or missing screening coverage information. Cases are separated by stage of disease (1 or 2) where this is known. Staging is based on parasitological detection in the cerebrospinal fluid or elevated white blood cell (WBC) count (>5 WBC/$\mu$l indicating stage 2 disease). Complete data used to fit the model are found in S1 Data and are visualised (without staging) here in Figs H–L alongside our model fits. We also show health districts with some data but not sufficient data points for fitting the model in S1 Data – our cut-off for model fitting was 10 data points where a single data point is either a year with active screening or a year with at least one passive case (e.g. if there are 5 years of data where there were passive cases each year and active screenings each year (with or without case detection) this is 10 data points).

Tsetse catch data from sentinel traps used to parameterise our tsetse dynamic model are discussed in the main text and presented in S2 Data.

Table A: **Population sizes and growth rates.**

| **Location** | **Population size (2014)** | **Population growth rate** |
| --- | --- | --- |
| Bonon (sub-prefecture) | 158,895 | 3.82% |
| Bouaflé (sub-prefecture) | 250,788 | 3.39% |
| Daloa (health district) | 591,633 | 2.00% |
| Oumé (health district) | 274,020 | 3.01% |
| Sinfra (health district) | 238,015 | 2.51% |

#### S1.2 The Compartmental gHAT Model

The gHAT model we considered in this study was the "Model 4" variant of the Warwick model previously presented in the literature [10, 19, 20, 14, 21]. This model variant includes high- and low-risk populations of people, with high-risk people not attending active screening while low-risk people attend active screening at random. gHAT infections among hosts are described by equation (S1.2.1) in our deterministic model version which was used for fitting to data. In the deterministic model each simulation using the same parameterisation yields the same model outcomes. We also utilise an analogous stochastic version of the model for estimating the year of elimination of transmission. This stochastic model has the same dynamics as the deterministic model for large population sizes and higher prevalences, but better captures dynamics of elimination around very low remaining case numbers. The corresponding event table for the stochastic model is found in Table B. In both deterministic and stochastic versions, human hosts are modelled by a susceptible-exposed-infectious-infectious-recovered-susceptible (SEIIRS) model with two infectious compartments, stage 1 disease, $I_{1H}$, and stage 2 disease, $I_{2H}$. Animal hosts that contribute to transmission are not included in this model, however animals that receive bites from tsetse but do not transmit are considered.

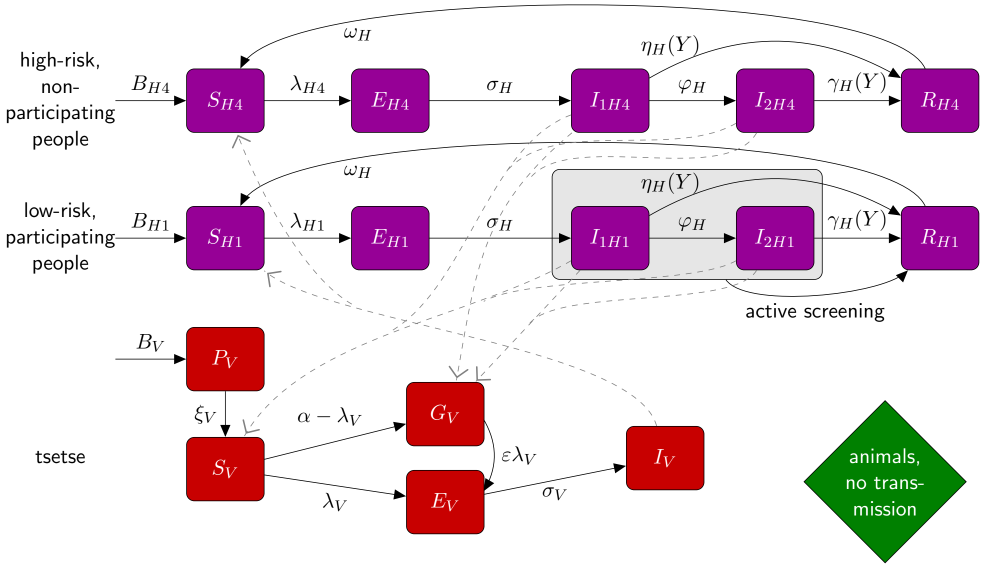


Fig A: **Schematic of compartmental model diagram.** The human population is partitioned into low-risk individuals (subscript $1$) who might participate in active screening, and high-risk individuals (subscript 4 – to maintain consistency with previously published variants of this model) who never participate in active screening. For each type of human subpopulation disease progression is the same, with individuals being born susceptible ($S_{Hi}$), and upon receiving an infectious bite from a infected tsetse they can become infected (exposed, $E_{Hi}$) and after their incubation period they develop stage 1 infection ($I_{1Hi}$), and if not detected and treated they progress to stage 2 infection ($I_{2Hi}$). Individuals who are detected in stage 1 or 2, or who die undetected move to the "recovered/removed" compartment. Tsetse natural history includes a pupal stage ($P_V$) and whilst unfed flies are fully susceptible to infection ($S_V$) we assume previously fed flies are less susceptible ($G_V$) – this is known as the teneral phenomenon. Although some tsetse blood feeding occurs on non-human animals, in this model variant, we assume no transmission cycle of the parasite occurs to and from tsetse and animals. This figure is adapted from [10] under a CC-BY licence.

Vectors are modelled by using compartments to appropriately model tsetse when used in a host-vector model with disease [20]. Pupal stage tsetse, $P_V$, emerge into unfed susceptible adults, $S_V$, and following a blood-meal become either exposed, $E_V$, or have reduced susceptibility to the *Trypanosoma brucei gambiense* parasites, $G_V$ - this effect is known as the teneral phenomenon. Following an infection, tsetse have an extrinsic incubation period (EIP) before becoming onwardly infectious. To incorporate a more realistic EIP distribution, there are three exposed classes, $E_{1V}, E_{2V}, E_{3V}$, which results in a gamma-distributed EIP (rather than an exponential EIP with a single exposed class).

In order to reduce the dimensionality of our ODE system (by one), the vector equations are non-dimensionalised using the scaling $N_H/N_V$, where $N_H$ is the total human population, and $N_V$ is the tsetse population size. This results in a new non-dimensionalised parameter, $m_{\text{eff}}$, which is $\frac{p_H N_V}{N_H}$ appearing in host equations ($p_H$ is the probability of a human being infected by a single infectious bloodmeal) and is referred to as the *effective vector density*.

The proportion of tsetse bites taken on low-risk and high-risk humans are $f_1$ and $f_4$, depending on the relative availability/attractiveness and the relative abundance of two risk groups. High-risk humans are assumed to be $r$-fold more likely to receive bites, i.e. $s_1 = 1$ and $s_4 = r$. Therefore, $f_i$'s can be calculated using $f_i = \frac{s_i N_{Hi}}{\sum_j s_j N_{Hj}}$.

$$
\text{Humans}\begin{cases}
\frac{dS_{Hi}}{dt} &= \mu_H N_{Hi} + \omega_H R_{Hi} - \alpha m_{\text{eff}} f_i \frac{S_{Hi}}{N_{Hi}} I_V - \mu_H S_{Hi}\\
\frac{dE_{Hi}}{dt} &= \alpha p_H f_i \frac{S_{Hi}}{N_{Hi}} I_V - (\sigma_H + \mu_H) E_{Hi}\\
\frac{dI_{1Hi}}{dt} &= \sigma_H E_{Hi} - (\varphi_H + \eta_H(Y) + \mu_H) I_{1Hi}\\
\frac{dI_{2Hi}}{dt} &= \varphi_H I_{1Hi} - (\gamma_H(Y) + \mu_H) I_{2Hi}\\
\frac{dR_{Hi}}{dt} &= \eta_H(Y) I_{1Hi} + \gamma_H(Y) I_{2Hi} - (\omega_H + \mu_H) R_{Hi}
\end{cases}
$$

$$
\text{Tsetse}\begin{cases}
\frac{dP_V}{dt} &= B_V N_H - (\xi_V + \frac{P_V}{K}) P_V\\
\frac{dS_V}{dt} &= \xi_V \mathbb{P}(\text{survive pupal stage}) P_V - \alpha S_V - \mu_V S_V\\
\frac{dE_{1V}}{dt} &= \alpha\big(1 - f_T(t)\big) p_V \big( \sum_i f_i \frac{(I_{1Hi} + I_{2Hi})}{N_{Hi}} + f_A \frac{I_A}{N_A}\big)(S_V + \varepsilon G_V)\\
&\quad -(3\sigma_V + \mu_V + \alpha f_T(t)) E_{1V}\\
\frac{dE_{2V}}{dt} &= 3\sigma_V E_{1V} - (3\sigma_V + \mu_V + \alpha f_T(t)) E_{2V}\\
\frac{dE_{3V}}{dt} &= 3\sigma_V E_{2V} - (3\sigma_V + \mu_V + \alpha f_T(t)) E_{3V}\\
\frac{dI_V}{dt} &= 3\sigma_V E_{3V} - (\mu_V + \alpha f_T(t)) I_V\\
\frac{dG_V}{dt} &= \alpha(1 - f_T(t))\Big(1 - p_V\big(\sum_i f_i \frac{(I_{1Hi} + I_{2Hi})}{N_{Hi}} + f_A \frac{I_A}{N_A}\big)\Big) S_V\\
&\quad -\alpha\Big(f_T(t) + (1 - f_T(t)) p_V \varepsilon\big(\sum_i f_i \frac{(I_{1Hi} + I_{2Hi})}{N_{Hi}} + f_A \frac{I_A}{N_A}\big)\Big) G_V\\
&\quad -\mu_V G_V
\end{cases}
\tag{S1.2.1}
$$

###### S1.2.0.1 Simulation of the model

As in previously presented studies, our ODE models were run assuming that, prior to 1998, they were at their endemic equilibrium. This endemic equilibrium was calculated analytically, based on model parameterisation, and used as an initial condition in the model code. In 1998 we assume active screening began at the same level reported in 2000 and that there was improvement to passive screening due to the availability of the CATT diagnostic test. In our model this has the effect of perturbing the dynamics away from endemic equilibrium and reducing transmission. Code for running deterministic and stochastic model version is available online (`https://osf.io/jtrs9/`).

**Deterministic version** The deterministic model ODEs were simulated between years using Matlab's "ode45" package (a Runge-Kutta (4,5) method), whilst active screening events at the beginning of years was applied as a discrete step change.

**Stochastic version** In order to permit a more accurate assessment of elimination of transmission by modelling, a discrete human population with discrete infection events was used and allowed us to identify the final transmission to humans. We choose to perform this via a tau-leap algorithm with a time step of one day, as per previous modelling studies by our group [11, 3]. The time step of one day was chosen due to the slow nature of gHAT, allowing the model to be simulated in a reasonable amount of time whilst still very closely approximating a direct simulation (i.e. via Gillespie's direct method). In order to do this, the equations modelling the human population were split into their constituent events as shown in Table B. Birth and death events were assumed to balance, and therefore avoid the need to create additional events for the two. At the start of each day, the rate at which any event is expected to occur was recalculated, and a Poisson sample was drawn with this rate, which was the number of events to be simulated as happening in this time period. The tsetse population was modelled deterministically using the Runge-Kutta 4th order method since we do not know the exact total population of tsetse, only a relative density.

| **Event** | **Effect** | | | | | | **Rate** |
|---|---|---|---|---|---|---|---|
| Become infected | $S_{H_i}$ | $\rightarrow$ | $S_{H_i}-1$ | $E_{H_i}$ | $\rightarrow$ | $E_{H_i}+1$ | $\alpha m_{\text{eff}} f_i \frac{S_{H_i}}{N_{H_i}} I_V$ |
| Become infectious | $E_{H_i}$ | $\rightarrow$ | $E_{H_i}-1$ | $I_{1H_i}$ | $\rightarrow$ | $I_{1H_i}+1$ | $\sigma_H E_{H_i}$ |
| Progress to stage 2 | $I_{1H_i}$ | $\rightarrow$ | $I_{1H_i}-1$ | $I_{2H_i}$ | $\rightarrow$ | $I_{2H_i}+1$ | $\varphi_H I_{1H_i}$ |
| Passive detection from stage 1 | $I_{1H_i}$ | $\rightarrow$ | $I_{1H_i}-1$ | $R_{H_i}$ | $\rightarrow$ | $R_{H_i}+1$ | $\eta_H(Y) I_{1H_i}$ |
| Passive detection from stage 2 or death | $I_{2H_i}$ | $\rightarrow$ | $I_{2H_i}-1$ | $R_{H_i}$ | $\rightarrow$ | $R_{H_i}+1$ | $\gamma_H(Y) I_{2H_i}$ |
| Die naturally whilst in $E_{H_i}$ | $E_{H_i}$ | $\rightarrow$ | $E_{H_i}-1$ | $S_{H_i}$ | $\rightarrow$ | $S_{H_i}+1$ | $\mu_H E_{H_i}$ |
| Die naturally whilst in $I_{1H_i}$ | $I_{1H_i}$ | $\rightarrow$ | $I_{1H_i}-1$ | $S_{H_i}$ | $\rightarrow$ | $S_{H_i}+1$ | $\mu_H I_{1H_i}$ |
| Die naturally whilst in $I_{2H_i}$ | $I_{2H_i}$ | $\rightarrow$ | $I_{2H_i}-1$ | $S_{H_i}$ | $\rightarrow$ | $S_{H_i}+1$ | $\mu_H I_{2H_i}$ |
| Die naturally whilst in $R_{H_i}$ or lose immunity | $R_{H_i}$ | $\rightarrow$ | $R_{H_i}-1$ | $S_{H_i}$ | $\rightarrow$ | $S_{H_i}+1$ | $(\mu_H+\omega_H) R_{H_i}$ |

Table B: **Events table for events affecting humans and their rates.** These rates are used in the stochastic version of the model

##### S1.2.1 Model parameterisation

The model takes various parameters. Some of them are fixed as they are not believed to vary geographically and there are estimate for these in the literature (see Table C). The remaining parameters are fitted in each different location based on reported case and screening data (see Table D). Sensitivity analysis on this model performed elsewhere [2] ranked these fitted parameters to have higher sensitivity compared to the fixed parameters using a ranked partial correlation method of this model variant.

Table C: **Model parameterisation (fixed parameters).** Notation, a brief description, and the values used for fixed parameters.

| Notation | Description | Value | |
|---|---|---|---|
| $N_H$* | Total human population size in 2015 | Fixed for each health zone | [16] |
| $\mu_H$ | Natural human mortality rate | $5.4795 \times 10^{-5}$ days$^{-1}$ | [23] |
| $B_H$ | Total human birth rate | $= \mu_H N_H$ | |
| $\sigma_H$ | Human incubation rate | 0.0833 days$^{-1}$ | [22] |
| $\varphi_H$ | Stage 1 to 2 progression rate | 0.0019 days$^{-1}$ | [5, 7] |
| $\omega_H$ | Recovery rate or waning-immunity rate | 0.006 days$^{-1}$ | [15] |
| Sens | Active screening diagnostic sensitivity | 0.91 | [6] |
| $B_V$† | Tsetse birth rate (per capita rate of depositing new pupae) | 0.0505 days$^{-1}$ | [20] |
| $\xi_V$ | Rate of pupal development to adult flies | 0.037 days $^{-1}$ | [20] |
| $K$‡ | Pupal carrying capacity | $= 111.09 N_H$ | [20] |
| $\mathbb{P}$(pupating) | Probability of a pupa surviving to emerge as an adult fly | 0.75 | [20] |
| $\mu_V$ | Tsetse mortality rate | 0.03 days$^{-1}$ | [22] |
| $\sigma_V$ | Tsetse incubation rate | 0.034 days$^{-1}$ | [12, 17] |
| $\alpha$ | Tsetse bite rate | 0.333 days$^{-1}$ | [26] |
| $p_V$ | Probability of tsetse infection per single infective bite | 0.065 | [22] |
| $\varepsilon$ | Reduced susceptibility factor for non-teneral (previously fed) flies | 0.05 | [19] |
| $f_H$ | Proportion of blood-meals on humans | 0.09 | [8] |
| $\eta_H^{\text{pre}}$ | Treatment rate from stage 1, pre-1998 | 0 | Assumed |
| $\text{disp}_{\text{act}}$§ | Overdispersion parameter for active detection | $4 \times 10^{-4}$ | [10] |
| $\text{disp}_{\text{pass}}$§ | Overdispersion parameter for passive detection | $2.8 \times 10^{-5}$ | [10] |

*The model is internally scaled such that the population size in all years corresponds to the population in 2014 (outputs are back-transformed to reflect focus-specific annual population growth rates).
†The value of $B_V$ was chosen to maintain constant population size in the absence of vector control interventions.
‡The value of $K$ was chosen to reflect the observed bounce back rate.
§Over-dispersion values were chosen based on previous model fitting work for DRC [10].

Table D: **Model parameterisation (fitted parameters).** Notation, brief description, and information on the prior distributions for fitted parameters.

| Notation | Description | Prior distribution* | Percentiles of prior distribution [2.5, 50 & 97.5%] | Unit |
|---|---|---|---|---|
| $R_0$ | Basic reproduction number (NGM approach) | $1 + \text{Exp}(10)$ | [1.003, 1.069, 1.369] | - |
| $r$ | Relative bites taken on high-risk humans | $1 + \Gamma(3.68, 1.09)$ | [2.015, 4.654, 10.028] | - |
| $k_1$ | Proportion of low-risk people | $\text{B}(16.97, 3.23)$ | [0.6564, 0.8514, 0.9609] | - |
| $\eta_H^{\text{post}}$ | Treatment rate from stage 1, 1998 onwards | $\Gamma(3.54, 5.32 \times 10^{-5})$ | [4.59, 17.1, 42.9]$\times 10^{-5}$ | days$^{-1}$ |
| $\gamma_H^{\text{post}}$ | Combined treatment and disease-induced death rate from stage 2, 1998 onwards | $\Gamma(6.2082, 0.001)$ | [2.33, 5.88, 12.0]$\times 10^{-3}$ | days$^{-1}$ |
| $\gamma_H^{\text{pre}}$ | Treatment and disease-induced death rate from stage 2, pre-1998 | $\Gamma(6.2082, 0.001)$ | [2.33, 5.88, 12.0]$\times 10^{-3}$ | days$^{-1}$ |
| Spec | Active screening diagnostic specificity | $0.998 + (1 - 0.998)\,\text{B}(7.23, 2.41)$ | [0.9989, 0.9995, 0.9999] | - |
| $u$ | Proportion of stage 2 cases reported from passive screening | $\text{B}(20, 40)$ | [0.2208, 0.3315, 0.4564] | - |
| $d_{\text{change}}$ | Midpoint year for passive improvement | $2000 + (2021 - 2000)\,\text{B}(47.53, 10.31)$ | [2015.0, 2017.3, 2019.1] | year |
| $\eta_{H_{\text{amp}}}$ | Relative improvement in passive screening stage 1 detection rate | $\Gamma(2, 5)$ | [1.21, 8.39, 27.86] | - |
| $\gamma_{H_{\text{amp}}}$ | Relative improvement in passive screening stage 2 detection rate | $\Gamma(2, 5)$ | [1.21, 8.39, 27.86] | - |
| $d_{\text{steep}}$ | Speed of improvement in passive screening detection rate | $\Gamma(15.71, 0.51)$ | [4.553, 7.843, 12.433] | years$^{-1}$ |

*Where Exp(.), $\Gamma(.)$ and B(.) are the exponential, gamma (parameterised with shape and scale) and beta distributions, respectively.

#### S1.3 Modelling passive detection and its improvement

Modelling passive detection and its improvement for Côte d'Ivoire follows the same methods as Crump et al. [10] which fitted to case data from the Democratic Republic of Congo and this is described below for completeness.

There are two sources of passive detection improvements considered in our model: a rapid improvement due to the introduction of the card agglutination test for trypanosomiasis (CATT) test in 1998 and a later improvement due to recent efforts to further improve the passive surveillance system, including training for health care workers and the introduction of rapid diagnostic tests as described in the main text. Prior distributions and percentiles of parameters related to passive detection and its improvement over time are summarised in Table E.

For improvements after 1998 we use the following equations to describe transmission rates from infected classes:

$$\eta_H(Y) = \eta_H^{\text{post}} \left[ 1 + \frac{\eta_{H_{\text{amp}}}}{1 + \exp\left(-d_{\mathsf{steep}}(Y - d_{\mathsf{change}})\right)} \right] \tag{S1.3.1}$$

$$\gamma_H(Y) = \gamma_H^{\text{post}} \left[ 1 + \frac{\gamma_{H_{\text{amp}}}}{1 + \exp\left(-d_{\mathsf{steep}}(Y - d_{\mathsf{change}})\right)} \right] \tag{S1.3.2}$$

We assume that all stage 1 cases are reported, but that some of the exits from stage 2 are due to death from gHAT disease. In 1998 the reporting probability for an exit from stage 2 is given by $u$, however as the exit rate from stage 2 increases this reporting probability does not stay constant, but increases (proportionally more people would be detected and treated with higher exit rates). When we compute reporting rates from stage 2 we therefore use the following:

$$\text{Death rate} = (1-u)\gamma_H^{\text{post}} \tag{S1.3.3}$$

$$\text{Stage 2 reporting incidence} = (\gamma_H(Y) - \text{Death rate})(I_{2H1} + I_{2H4}) \tag{S1.3.4}$$

We considered improvement in passive screening systems over time for all foci and note that such improvements have been noted for other gHAT-endemic countries: firstly there have been on-the-ground changes to the tools used and training in passive screening which should enable more rapid detection, but data also support this hypothesis of improved passive detection with increasing proportions of passive cases being stage 1 infections [4].

Table E: **Parameterisation of passive detection improvement.** Notation and brief description of fitted parameters related to passive detection improvement plus their within former province prior distributions and [2.5th, 50th & 97.5th] percentile.

| **Parameter** | | | |
|---|---|---|---|
| | **Health district** | **Prior distribution** | **Percentiles of prior distribution** |
| $\eta_H^{\text{post}}$ – Treatment rate from stage 1, 1998 onwards | | | |
| | **All** | $\Gamma(3.54, 5.32 \times 10^{-5})$ | $[4.59, 17.1, 42.9] \times 10^{-5}$ |
| $\gamma_H^{\text{post}}$ – Treatment rate from stage 2, 1998 onwards | | | |
| | **All** | $\Gamma(2.45, 1.92 \times 10^{-3})$ | $[7.59, 40.7, 121] \times 10^{-4}$ |
| $\eta_{H_{\text{amp}}}$ – Relative improvement in passive stage 1 detection rate | | | |
| | **With improvement** | $\Gamma(2, 5)$ | $[1.21, 8.39, 27.86]$ |
| $\gamma_{H_{\text{amp}}}$ – Relative improvement in passive stage 2 detection rate | | | |
| | **With improvement** | $\Gamma(2, 5)$ | $[1.21, 8.39, 27.86]$ |
| $d_{\mathsf{steep}}$ – Speed of improvement in passive detection rate | | | |
| | **With improvement** | $\Gamma(39.6, 2.70 \times 10^{-2})$ | $[0.761, 1.06, 1.42]$ |

#### S1.4 Modelling vector control

In the present study we utilise the same method to simulate the impact of annual vector control on tsetse populations as presented elsewhere [20, 10, 13]. We model the dynamics of tsetse populations in the presence of Tiny Target-based vector control on the basis of an assumed tsetse population density reduction measured by the entomological monitoring trap catches pre- and post-deployment presented in the main text. We note that the reductions are very high, but not dissimilar to reductions presented elsewhere using this technology [9, 24, 25, 14]. Fig B shows the modelled dynamics of tsetse populations in the model (blue line).

We utilise a function which describes the probability of a host-seeking tsetse both hitting a Tiny Target and dying as a result, $f_T$, is time dependent ($t$, in days, measured from when the targets were deployed):

$$f_T(t) = f_{\text{max}} \left(1 - \frac{1}{1 + \exp(-0.068(\text{mod}(t, 365) - 127.75))}\right) \tag{S1.4.1}$$

and $f_{\text{max}}$ - the maximum daily probability of contacting a Tiny Target and dying as a result. $f_T$ modifies all the bite rates $\alpha$ in our tsetse equations to produce an additional Tiny-Target-induced mortality for tsetse.

The effectiveness of Tiny Targets is assumed to wane over time so that at six months after deployment, the targets are virtually non-effective. To simulate this we have a rapid decrease in effectiveness after 127.75 days, determined by the value -0.068, after this time [20]. This represents lost Tiny Targets (e.g. due to rainfall) or loss of the effectiveness of remaining targets (e.g. vegetation growth impacting their visibility). Here we used maximum likelihood estimation to fit the model parameters $\theta = (f_{\text{max}}, B_V)$ to the trap data in Bonon and Sinfra foci (see S2 Data). The parameter $K$ was adjusted so that the equilibrium density of adult tsetse before control was constant, $N_V^*$. We assumed that the number of tsetse observed in each survey was Poisson distributed about the level predicted by the model. Denoting the modelled tsetse density at time $t$ as $g(t)$, and the data as $(t_i, n_i)$, the likelihood function is given by:

$$\begin{aligned} LL(\theta, g(0)|\text{VC data}) &= \log(\mathbb{P}(\text{VC data}|\theta, g(t_0))) \\ &= \log\left(\prod_i \mathbb{P}(n_i = g(t_i)|\theta, g(t_0))\right) \\ &= \sum_i \log\left(\mathbb{P}(n_i = g(t_i)|\theta, g(t_0))\right) \\ &= \sum_i \log\left(\frac{g(t_i)^{n_i} e^{-g(t_i)}}{n_i!}\right) \\ &= \sum_i \left(n_i \log g(t_i) - g(t_i) - \log n_i!\right) \end{aligned} \tag{S1.4.2}$$

The best model fits to the catch data, $g(t)$, and functions $f_T(t)$ for each focus are shown in Fig B. Fitted parameter values for each focus are found in Table F. In Sinfra, the first deployment was in May 2017, whereas all future deployments were in July.

Table F: **Maximum likelihood estimates for the tsetse population model parameters**

| **Parameter** | **Bonon** | **Sinfra** |
|---|---|---|
| $f_{\text{max}}$ | 0.2138 | 0.1347 |
| $B_V$ | $6.199 \times 10^{-2}$ | $5.347 \times 10^{-2}$ |

Although this study does not focus on future projections we do show what the future modelled dynamics would be assuming deployments of targets stopped after 2021 (Fig B dashed lines in green and orange) or continued after 2021 (Fig B solid lines in green and orange). This is shown to demonstrate that our tsetse model is capable of representing possible bounceback of tsetse populations following cessation of target deployments and the speed at which the model predicts this would happen.

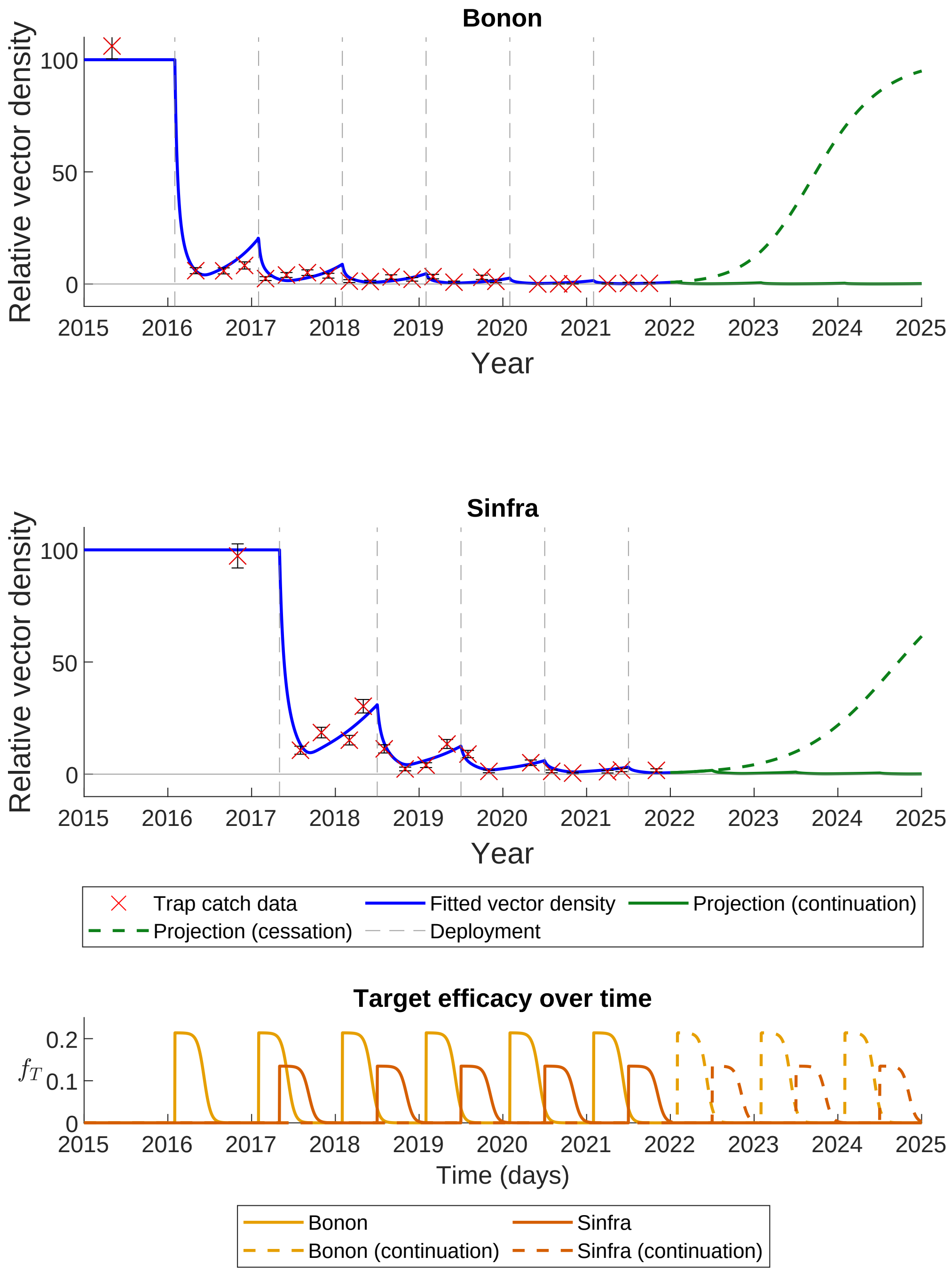


Fig B: **Impact of Tiny Targets on tsetse density.** The figures show how varying target efficacy (lower plot) impacts tsetse population density (upper plots). Target efficacy is measured as the proportion of a host-seeking tsetse which will both hit the Tiny Target and die as a result. The graphs show the necessary efficacy of targets needed to provide the best fit (maximum likelihood estimate) for Bonon (top) and Sinfra (bottom) to the entomological monitoring data shown as red crosses. From 2022 onwards we show two options (i) continuation of further target deployments (solid lines), or (ii) cessation of the vector control intervention (dashed lines).

#### S1.5 Fitting

##### S1.5.1 Running the models

For fitting the deterministic model, there are two elements, each of which is initialised.

- Two chains are run in the Markov chain Monte Carlo (MCMC) used in the fitting. The chains are initialised using the fixed parameters and by random perturbations around supplied, individually valid, initial values of each parameter being fitted, rejecting those parameter sets that do not produce a valid posterior probability.
- Each projection is initialised with a randomly sampled realisation from the posterior distribution of fitted parameters alongside the set of fixed parameters.

##### S1.5.2 Likelihood

12 parameters; $R_0, r, \eta_H, \gamma_H, \gamma_{H0}, k_1, u$, and Spec, $d_{\text{change}}$, $\eta_{H_{\text{amp}}}$, $\gamma_{H_{\text{amp}}}$, $d_{\text{steep}}$ were fitted in all foci.

For fitting the model to case data we transform model ODE solutions (for S1.2.1) into annual case reporting denoted $A_{M1}$, $A_{M2}$, for active stage 1 and stage 2 and $P_{M1}$, $P_{M2}$, for passive stage 1 and 2. Since we always know the stage (1 or 2) in the model simulations there is no requirement for a "U" (unknown stage) category for the model. These are computed using solutions to the ODEs for the given set of parameters aggregated across a year.

In terms of main text Fig 1, they relate to the transfer from infectious categories to the recovered category – the new annual reported case incidence. This is either by passive detection from stage 1 for year $Y$:

$$P_{M1}(Y) = \int_{Y}^{Y+1} \eta_H(Y)\big(I_{1H1}(t) + I_{1H4}(t)\big)\,\mathrm{d}t,$$

passive detection from stage 2

$$P_{M2}(Y) = \int_{Y}^{Y+1} (\gamma_H(Y) - \text{Death rate})\big((I_{2H1}(t) + I_{2H4}(t)\big)\,\mathrm{d}t,$$

or by active screening from the low-risk $(H1)$ group in year $Y$

$$A_{M1}(Y) = z(Y)\text{Sens}I_{1H1}(Y) + z(Y)(1 - \text{Spec})\big(k_1 N_H - I_{1H1}(Y) - I_{2H1}(Y)\big)$$

and

$$A_{M2}(Y) = z(Y) \times \text{Sens} \times I_{2H1}(Y)$$

with variable active screening coverage by year, $z(Y)$ and fixed diagnostic sensitivity. $A_{M1}$ also contains any false positives that may have been incorrectly identified from non-infected people based on the high but imperfect specificity of the active screening algorithm. We assume in Côte d'Ivoire that all false positives would be assigned to be stage 1 and treated, however in the model false positives stay in the susceptible class unlike true cases which move to recovereds.

The log-likelihood function used in the adaptive Metropolis-Hastings MCMC contained two terms in each year for which reported case numbers were available for each source of reported cases (active or passive screening). These were:

- a beta-binomial probability that the total number of cases reported in that year for that source came from the available population (either the reported number of people actively screened for active screening, or the health zone population for passive screening) with probability calculated from solving the ODE for the current set of parameters, and
- a binomial probability that the reported stage 1 cases come from the total number of reported staged cases where the probability parameter again comes from the solution of the ODE. In many years staging is unknown and so this part of the log-likelihood will return zero and not contribute to our calculation. In some years, we only partially know staging information.

This formulation allowed over-dispersion in the observed cases to be included, via the beta-binomial distribution, and any proportion of cases with reported disease stage to be appropriately accounted for (assuming that the reporting of staging information is independent of the disease stage). The log-likelihood function was as follows:

$$
\begin{aligned}
LL(\theta|x) = & \log(P(x|\theta)) \\
\propto & \sum_{i=2000}^{2016} \left( \log\left[ \text{BetaBin}\left( A_{D1}(i) + A_{D2}(i) + A_{DU}(i); z(i), \frac{A_{M1}(i) + A_{M2}(i)}{z(i)}, \text{disp}_{\text{act}} \right) \right] \right. \\
& + \log\left[ \text{Bin}\left( A_{D1}(i); A_{D1}(i) + A_{D2}(i), \frac{A_{M1}(i)}{A_{M1}(i) + A_{M2}(i)} \right) \right] \\
& + \log\left[ \text{BetaBin}\left( P_{D1}(i) + P_{D2}(i) + P_{DU}(i); N_H, \frac{P_{M1}(i) + P_{M2}(i)}{N_H}, \text{disp}_{\text{pass}} \right) \right] \\
& \left. + \log\left[ \text{Bin}\left( P_{D1}(i); P_{D1}(i) + P_{D2}(i), \frac{P_{M1}(i)}{P_{M1}(i) + P_{M2}(i)} \right) \right] \right)
\end{aligned}
$$

The model takes parameterisation $\theta$, $x$ is the data, $P_{Dj}(i)$ and $A_{Dj}(i)$ are the number of cases detected by passive or active screening (of stage $j$, which may be 1, 2 or unknown, $U$) in year $i$ of the data. $P_{Mj}(i)$ and $A_{Mj}(i)$ are the number of cases detected by passive or active screening (of stage $j$) in year $i$ of the model, and $z(i)$ is the number of people actively screened in year $i$. $\text{BetaBin}(m; n, p, \rho)$ gives the probability of obtaining $m$ successes out of $n$ trials with probability $p$ and overdispersion parameter $\rho$. The overdispersion accounts for larger variance than under the binomial. The probability density function of this distribution is given by:

$$
\text{BetaBin}(m; n, p, \rho) = \frac{\Gamma(n+1)\Gamma(m+a)\Gamma(n-m+b)\Gamma(a+b)}{\Gamma(n-m+1)\Gamma(n+a+b)\Gamma(a)\Gamma(b)}
$$

where $a = p(1/\rho - 1)$ and $b = a(1-p)/p$.

##### S1.5.3 Imputation of missing numbers screened information

There are instances in the data where the number of cases from active screening within year $t$ ($A_D(t) = A_{D1} + A_{D2}$) is not consistent with the number of people recorded as having been screened in that year for that health zone ($z(t)$), i.e. $A_D(t) > z(t)$. In this situation, if $A_D(t) < 20$ we assume that these people have attended a screening outside their home focus and the record has been correctly allocated to their home focus and we set $z(t) = A_D(t)$. However, where $A_D(t) \geq 20$ we assume that a screening must have taken place in the focus but that only the positive test results have been recorded. To allow use of these records we impute the missing negative test results ($A_D^-(t)$) during the MCMC analysis, and this enables the model to account for perturbation of the system by unknown high levels of active screening activity within a year.

We use a geometric prior for $A_D^-(t)$; $A_D^-(t) \sim \text{Geom}(\lambda_t)$, where:

$$
\lambda_t = \frac{1}{1 + \bar{z}_t}, \qquad \bar{z}_t = \frac{\sum_{t^*=2000, t^* \neq t}^{2021} z_{t^*} e^{-|t-t^*|}}{\sum_{t^*=2000, t^* \neq t}^{2021} e^{-|t-t^*|}}
$$

such that $\bar{z}_t$ is a weighted average of numbers screened per year with higher weight given to years closer to $t$. The proposal distribution from which $A_D^-(t)$ is sampled is $A_D^-(t)|X_t, p_t \sim \text{NB}(X_t + 1, 1 - (1 - p_t)(1 - \lambda_t))$, where $p_t$ is the probability of a positive screening test result obtained from solutions of the ODE up to this point in time. The value $\hat{z}(t) = A_{D1} + A_{D2} + A_D^-(t)$ is used in place of the unknown $z(t)$ in the likelihood calculation.

Table G: **Imputation of negative active screening results.** Focus, year and case numbers from active screening ($A_D(t)$) where imputation of $A_D^-(t)$ was performed; plus median and 95% credible intervals of posterior distributions of $A_D^-(t)$.

| Health district/subprefecture | Year ($t$) | $A_D(t)$ | $A_D^-(t)$ | |
|---|---|---|---|---|
| | | | Median | [95% CI] |
| Bouaflé (Bonon) | 2001 | 11 | 7142 | [3658,15397] |
| | 2005 | 3 | 3054 | [1092,7840] |
| | 2014 | 1 | 1565 | [314,4862] |
| Oumé | 2000 | 3 | 4330 | [1638,10643] |
| Sinfra | 2001 | 1 | 1097 | [241,3157] |
| | 2004 | 6 | 5589 | [2696,11648] |
| | 2005 | 1 | 1165 | [218,3537] |
| | 2008 | 1 | 1161 | [244,3362] |
| | 2010 | 1 | 1125 | [265,3423] |

### S1.6 Additional results

#### S1.6.1 Posterior parameter distributions for all foci

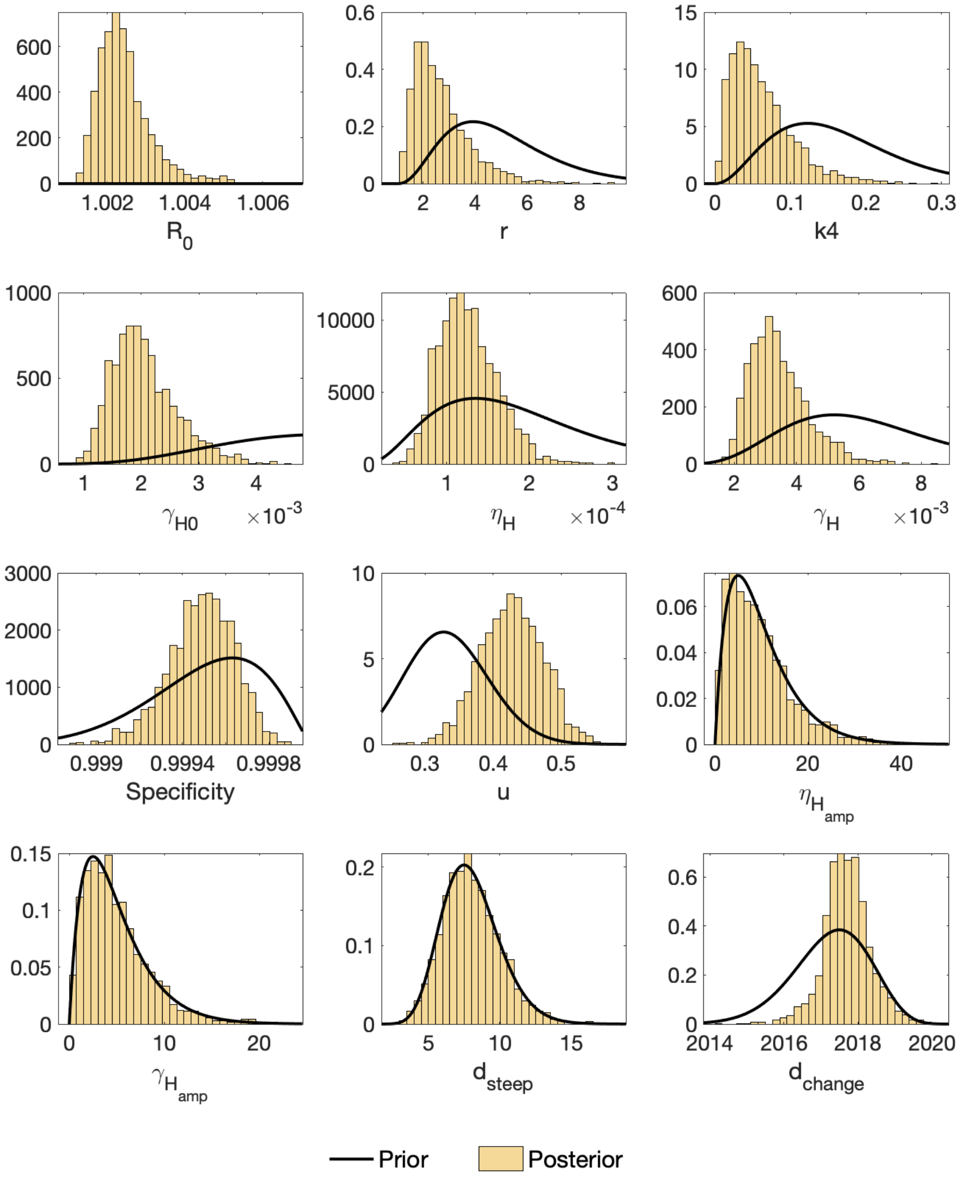


Fig C: **Posterior parameter distributions for Bonon subprefecture (of Bouaflé health district).** Histograms show the posterior parameter distributions after fitting the model (yellow). Black lines show the prior parameter distributions.

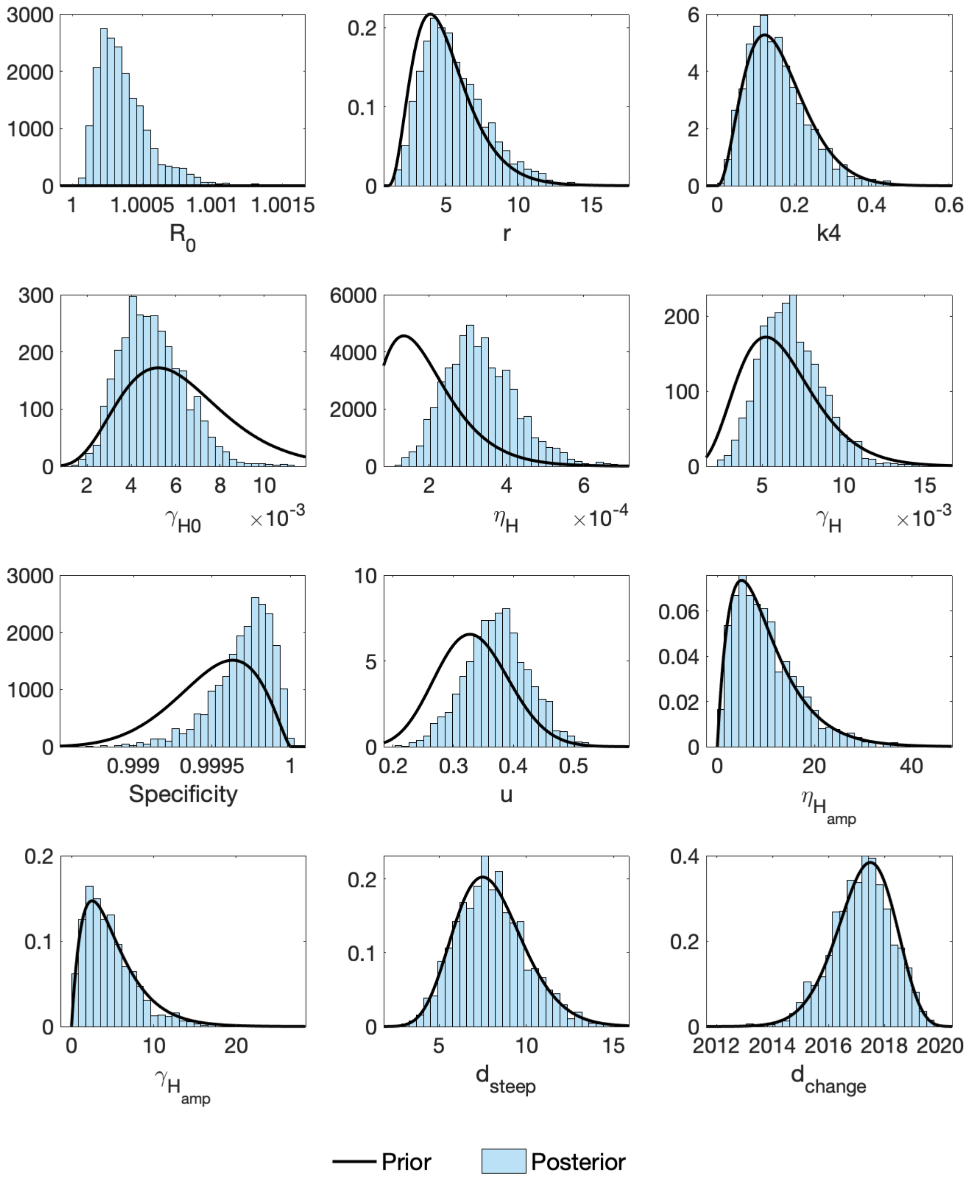


Fig D: **Posterior parameter distributions for Bouaflé subprefecture (of Bouaflé health district).** Histograms show the posterior parameter distributions after fitting the model (blue). Black lines show the prior parameter distributions.

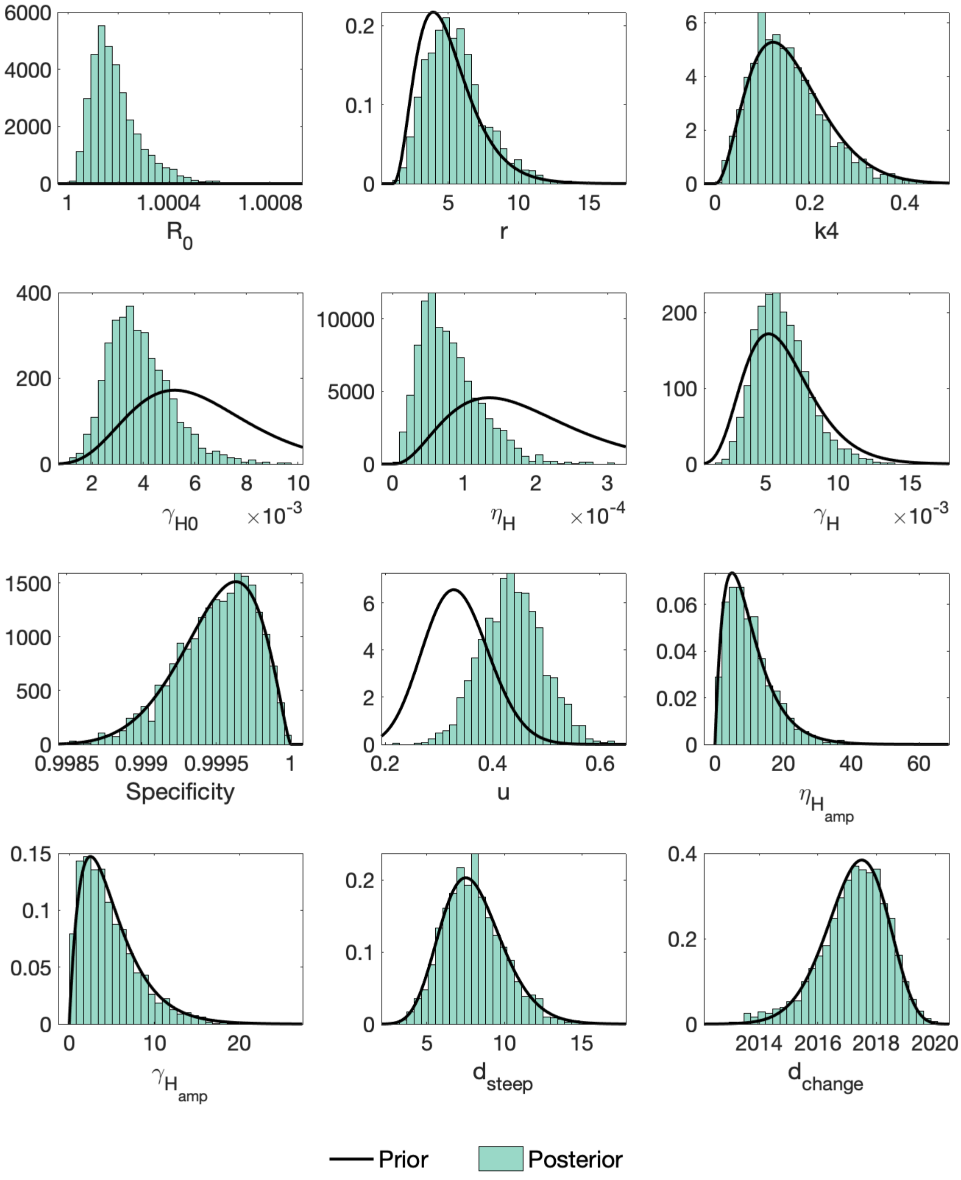


Fig E: **Posterior parameter distributions for Daloa health district.** Histograms show the posterior parameter distributions after fitting the model (green). Black lines show the prior parameter distributions.

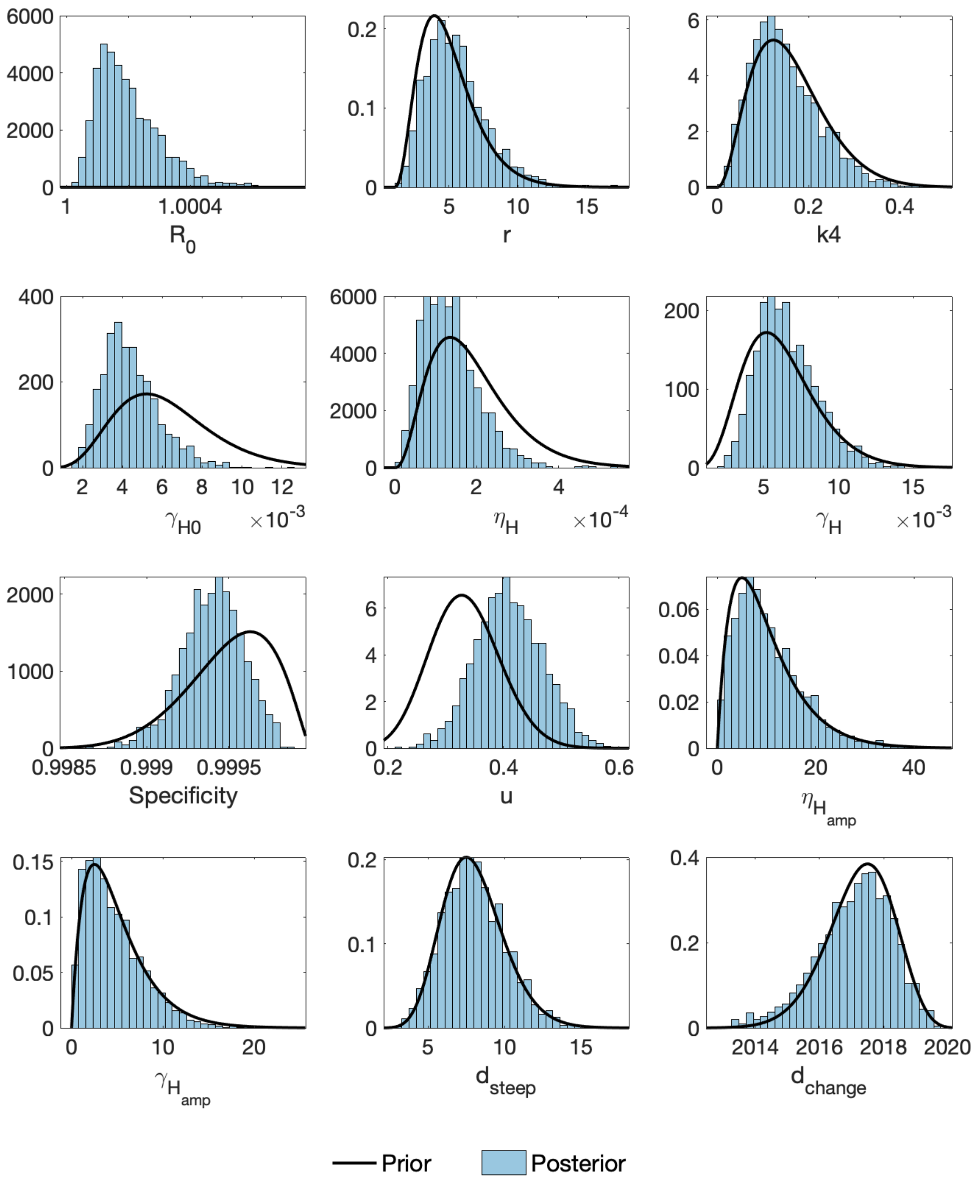


Fig F: **Posterior parameter distributions for Oumé health district.** Histograms show the posterior parameter distributions after fitting the model (blue). Black lines show the prior parameter distributions.

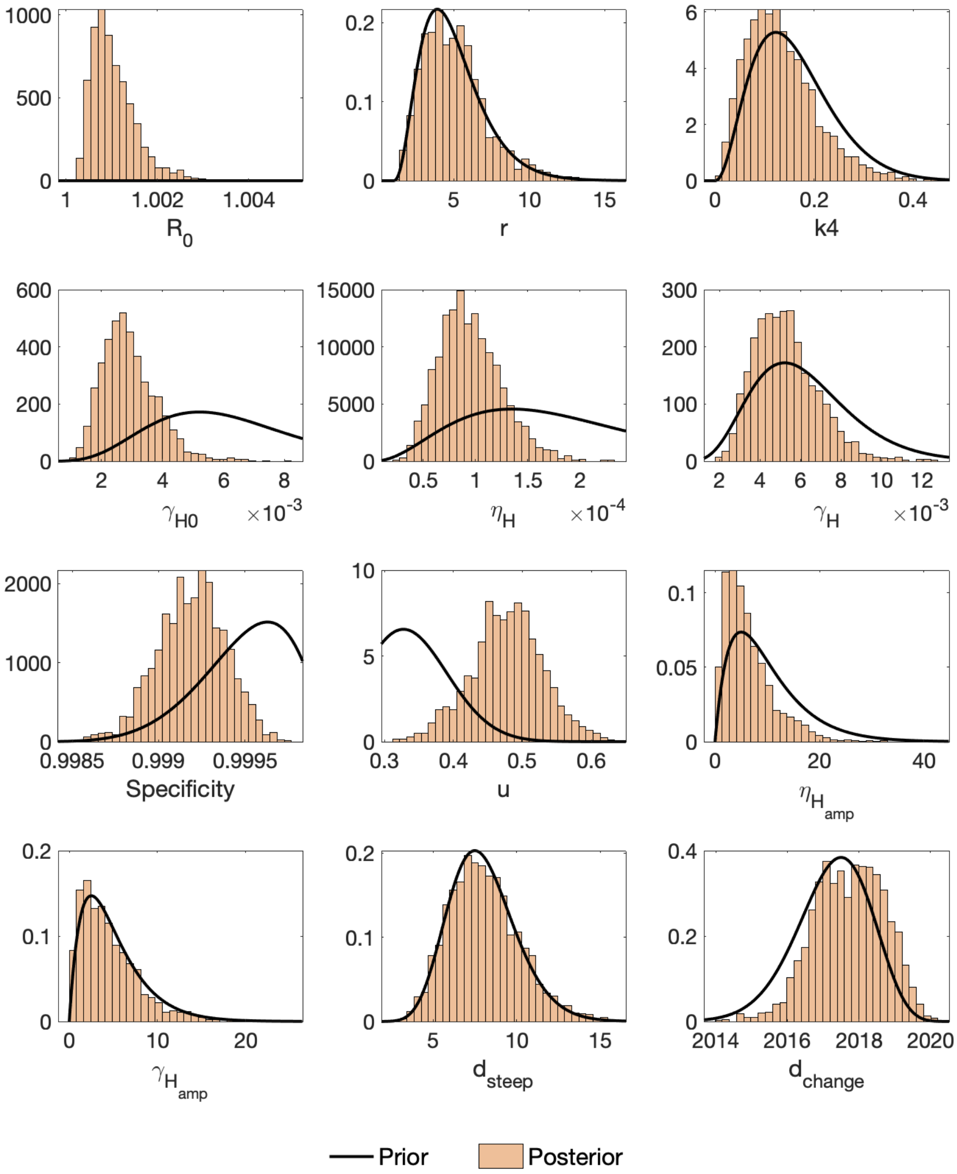


Fig G: **Posterior parameter distributions for Sinfra health district.** Histograms show the posterior parameter distributions after fitting the model (orange). Black lines show the prior parameter distributions.

##### S1.6.2 Reduction in transmission

Table H: **Estimated percentage reduction in historical transmission.** Deterministic model results were used to calculate this reduction for the three time periods (i) 2000 to 2010, (ii) 2011 to 2021 and (iii) 2000 to 2021. Medians and 95% credible intervals (CIs) are given.

| Focus | 2000–2010 Median [95% CI] | | 2011–2021 Median [95% CI] | | 2000–2021 Median [95% CI] | |
|---|---|---|---|---|---|---|
| Bouaflé | 70.9 | [60.0–79.6] | 100.0 | [100.0–100.0] | 100.0 | [100.0–100.0] |
| Bonon[1] | 71.9 | [58.9–81.4] | 100.0 | [100.0–100.0] | 100.0 | [100.0–100.0] |
| Bouaflé[1] | 67.4 | [42.9–82.9] | 100.0 | [100.0–100.0] | 100.0 | [100.0–100.0] |
| Daloa | 54.9 | [24.4–73.9] | 100.0 | [61.1–100.0] | 100.0 | [74.9–100.0] |
| Oume | 52.0 | [20.6–73.5] | 100.0 | [62.8–100.0] | 100.0 | [75.4–100.0] |
| Sinfra | 67.0 | [47.8–80.0] | 100.0 | [100.0–100.0] | 100.0 | [100.0–100.0] |
| Aggregate[2] | 66.1 | [57.9–73.1] | 100.0 | [89.6–100.0] | 100.0 | [96.5–100.0] |

[1]Subprefectures of Bouaflé district.

[2]New infections aggregated across the five independent analyses.

##### S1.6.3 Fits for all foci

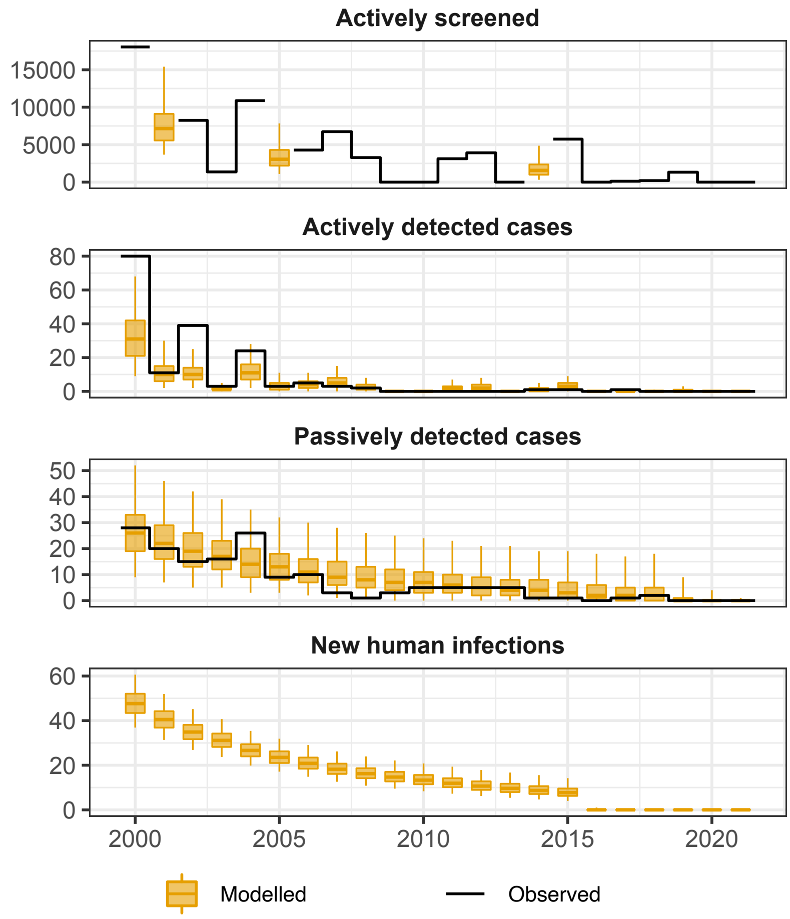


Fig H: **Model fit to 2000–2021 gHAT case data from Bonon subprefecture (of Bouaflé health district).** The black lines show the observed data (either number of people screened or cases) and the yellow box and whisker plots show the model (centre line is the median, boxes contain 50% credible intervals (CIs) and whiskers show 95% CIs). Some years of data are missing for total number of people actively screened so this was estimated during fitting with results shown as box and whiskers. New infections are not directly observable and are estimated through the model based on case reporting.

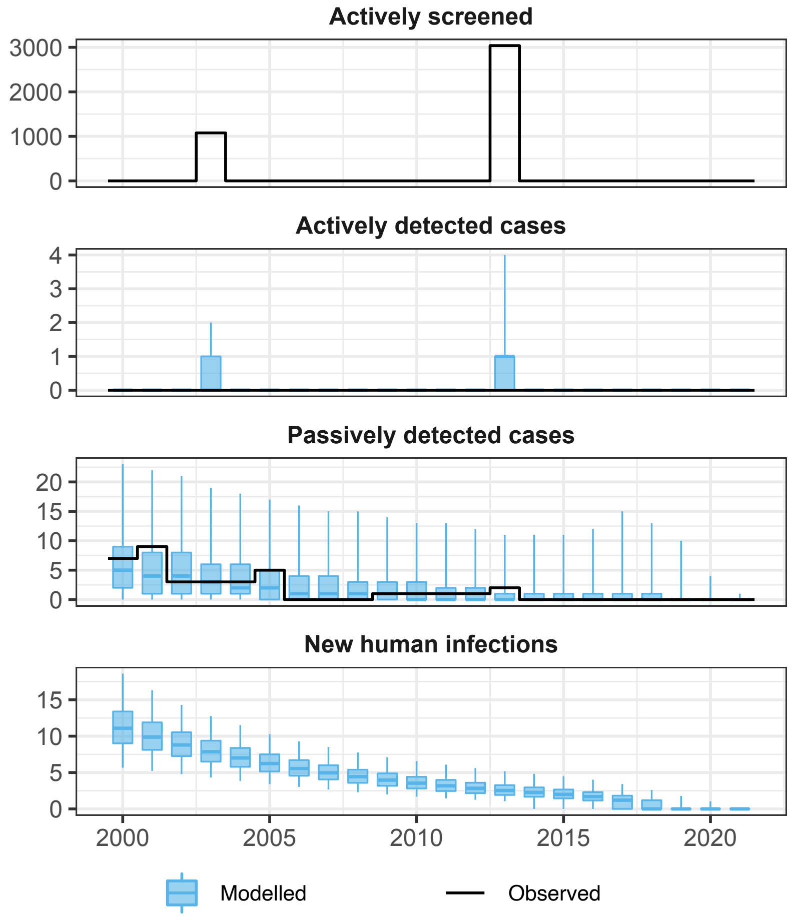


Fig I: **Model fit to 2000–2021 gHAT case data from Bouaflé subprefecture (of Bouaflé health district).** The black lines show the observed data (either number of people screened or cases) and the blue box and whisker plots show the model (centre line is the median, boxes contain 50% credible intervals (CIs) and whiskers show 95% CIs). Some years of data are missing for total number of people actively screened so this was estimated during fitting with results shown as box and whiskers. New infections are not directly observable and are estimated through the model based on case reporting.

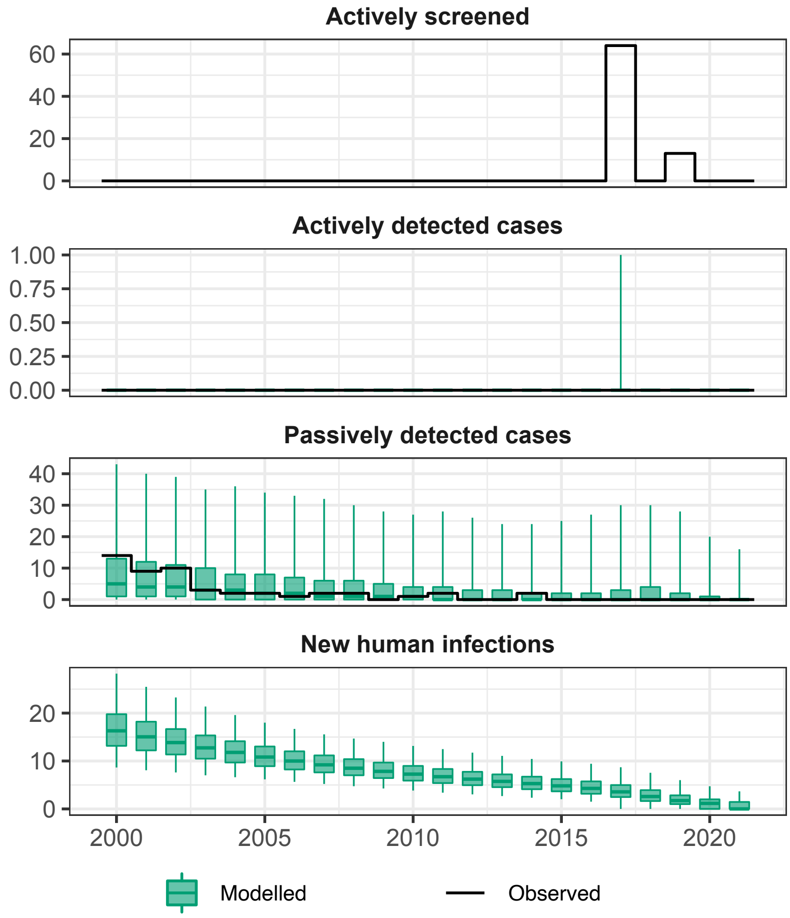


Fig J: **Model fit to 2000–2021 gHAT case data from Daloa health district.** The black lines show the observed data (either number of people screened or cases) and the green box and whisker plots show the model (centre line is the median, boxes contain 50% credible intervals (CIs) and whiskers show 95% CIs). Some years of data are missing for total number of people actively screened so this was estimated during fitting with results shown as box and whiskers. New infections are not directly observable and are estimated through the model based on case reporting.

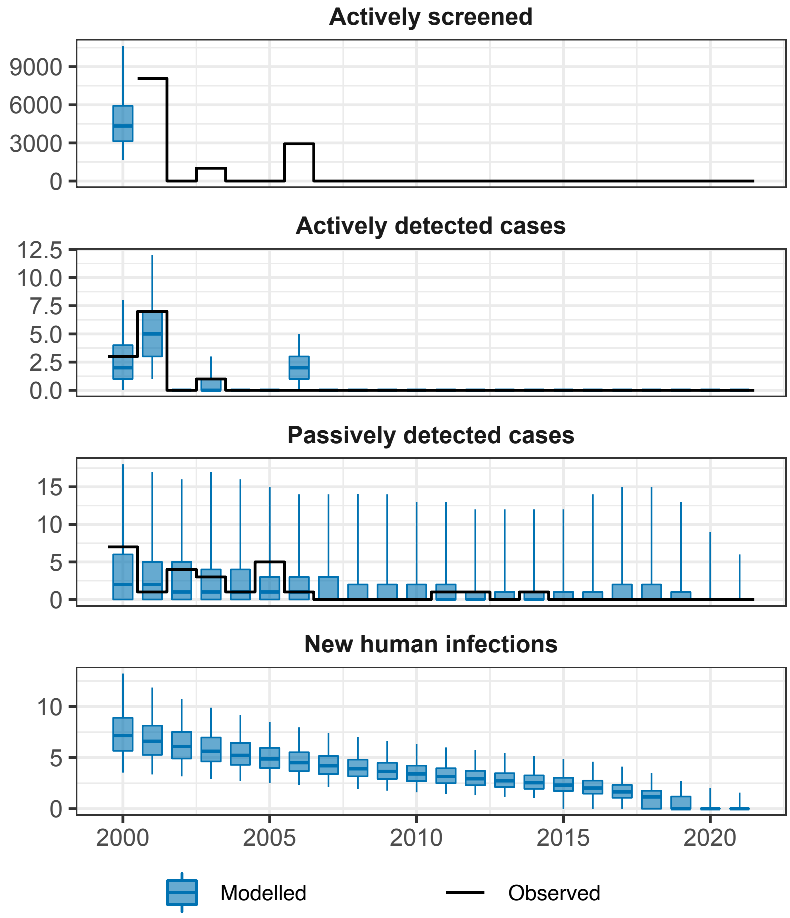


Fig K: **Model fit to 2000–2021 gHAT case data from Oumé health district.** The black lines show the observed data (either number of people screened or cases) and the blue box and whisker plots show the model (centre line is the median, boxes contain 50% credible intervals (CIs) and whiskers show 95% CIs). Some years of data are missing for total number of people actively screened so this was estimated during fitting with results shown as box and whiskers. New infections are not directly observable and are estimated through the model based on case reporting.

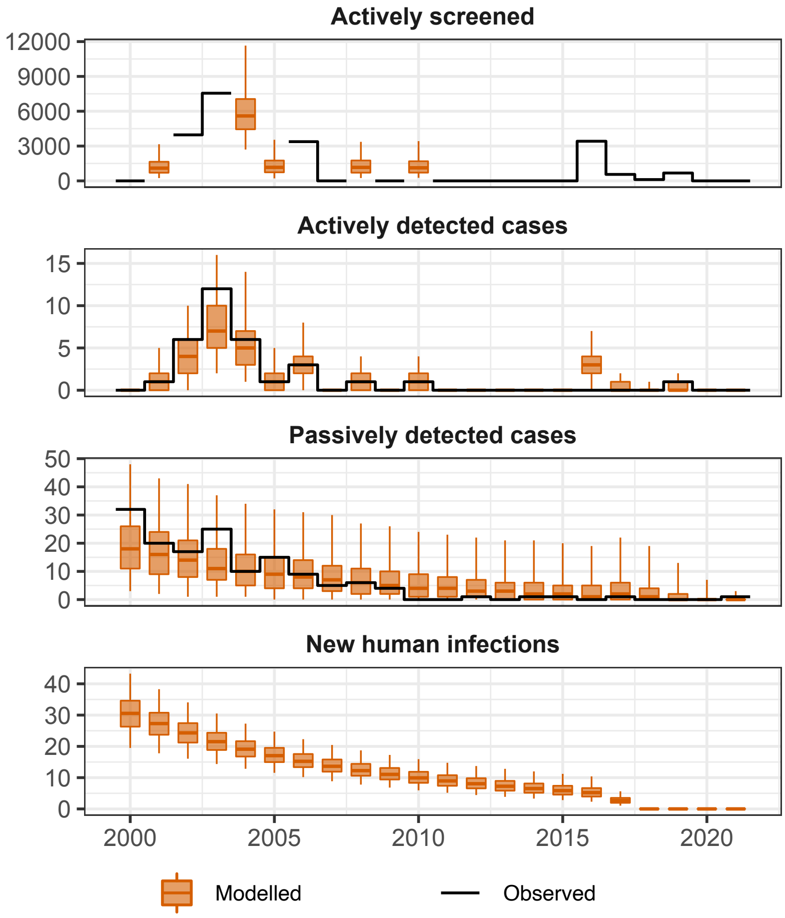


Fig L: **Model fit to 2000–2021 gHAT case data from Sinfra health district.** The black lines show the observed data (either number of people screened or cases) and the orange box and whisker plots show the model (centre line is the median, boxes contain 50% credible intervals (CIs) and whiskers show 95% CIs). Some years of data are missing for total number of people actively screened so this was estimated during fitting with results shown as box and whiskers. New infections are not directly observable and are estimated through the model based on case reporting.

##### S1.6.4 Estimating focus-level EoT

To estimate the probability of EoT we ran our analogous stochastic model using the posterior parameterisation found for each location through fitting the deterministic model. If we had used the deterministic model directly there would have been a need to define a proxy threshold to determine that EoT had been met. This problem, which arises as the deterministic model outputs for new infection can take non-integer values which asymptote but never attain zero new infections per year, has been described before elsewhere [1, 4, 13]. Use of the stochastic model overcomes this issue as it counts integer numbers of transmission events occurring each year.

We note that there is a difference between EoT and elimination of infection (EoI) and last case reporting. EoT must occur in the same year or earlier than EoI since the final infected person must recover or die after the last transmission event. The last case reporting can also occur at the same time as the last recovery or death (EoI) or before hand due to imperfect reporting.

In the model we can only know if the a transmission event is the last one once there is no remaining infection in the system (i.e. no $E_H, I_H$) however to limit our simulations to fixed time horizons we compute our EoT curves in the main text Figure 8 (for health districts) and here in Figure M (for disease foci) by using a cut off in 2052 – for any given year, $Y$, we say that there is EoT for a given realisation if there are no more transmission events during the period $[Y, 2052]$. This allows more than sufficient time to catch the vast majority of possible resurgence. In practise there is very little difference between this cut-off and waiting for EoI to reach zero for this gHAT model.

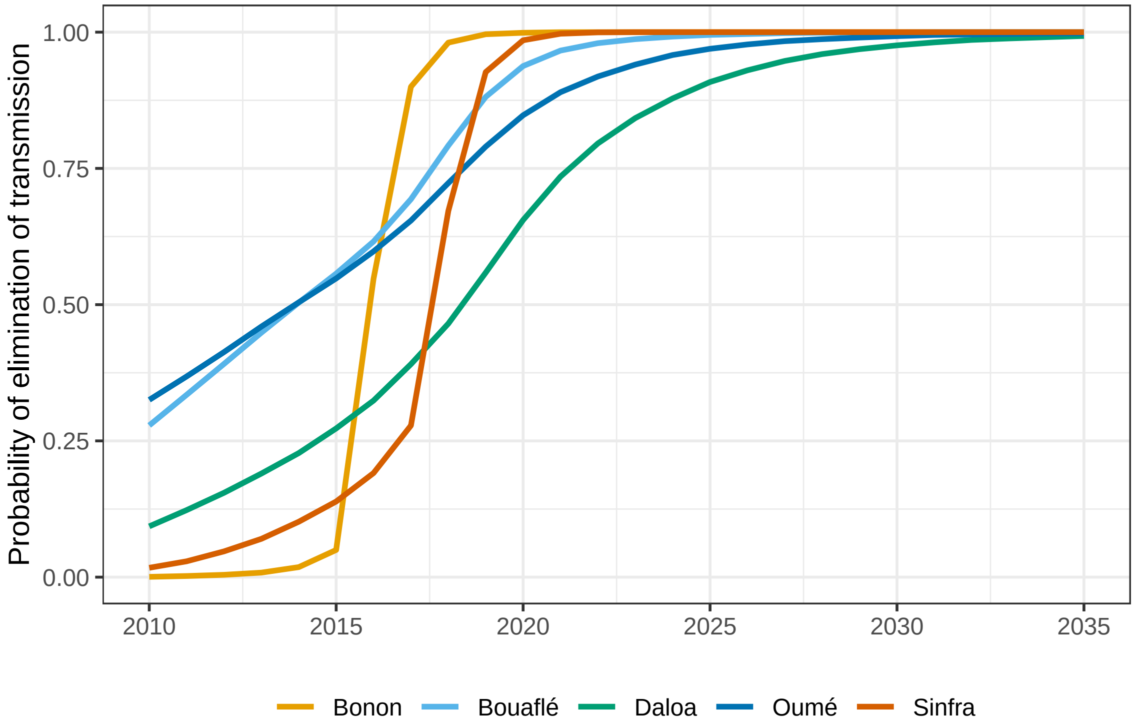


Fig M: **Estimated probability of elimination of transmission (EoT) by year for each fitted focus.** Daloa, Oumé and Sinfra are health districts, whilst Bonon and Bouaflé are sub-prefectures. The probability is computed by assessing the proportion of model simulations that fall below our proxy EoT threshold (< 1 new infection per year) for each year for each location

#### S1.7 PRIME-NTD criteria

Table I: PRIME-NTD criteria fulfillment. How the NTD Modelling Consortium's "5 key principles of good modelling practice" have been met in the present study.

| Principle | What has been done to satisfy the principle? | Where in the manuscript is this described? |
|---|---|---|
| **1. Stakeholder engagement** | | |
| | This study is one in a series of modelling analyses using real-world data from gHAT endemic countries, although this is the first for Côte d'Ivoire. This study was lead by members of the national sleeping sickness control programme in Côte d'Ivoire (PNETHA), Institut Pierre Richet, Université Jean Korougnon Guédé, and the Projet de Rechereches Clinques sur la Trypanosomiasis – coauthors Lingué Kouakou, Dramane Kaba, Mathurin Koffi, Emmanuel Kouassi N'Gouan, Vincent Djohan, Martial Kassi N'Djetchi, Bamoro Coulibaly, Djakaridja Berté, Bi Tra Dieudonné Ta, and Minayégninrin Koné – and their partners. The modelling team was invited to lead the simulation and analysis work guided by numerous in-person visits, online meetings and emails. These meetings have enabled the modellers to understand the context of how the human case data used in the study were collected, how interventions have been implemented across different geographies, and how this has changed over time. | Authorship list |
| **2. Complete model documentation** | | |
| | Full model fitting code and documentation is available through OpenScienceFramework (OSF). The model is fully described in the main text and SI. | See Materials and Methods section in the main text, Supplementary Information (file S1 Text) and at OSF (`https://osf.io/jtrs9/`) |
| **3. Complete description of data used** | | |
| | Aggregate data used for fitting are shown alongside model fits for each disease focus in the SI. S1 Data contains the case and screening data used in the MCMC. | See Figs H–L and S1 Data. |
| **4. Communicating uncertainty** | | |
| | *Structural uncertainty*: We used a previously developed model variant which includes heterogeneity in human risk to tsetse bites and has been shown to capture gHAT dynamics well in other settings [19, 21]. | *Structural uncertainty*: See section S1.3 in this SI. |
| | *Parameter uncertainty*: Parameter distributions are estimated by fitting to data. All model fits and projections include and propagate parameter uncertainty and include it in visual representations (either box and whisker plots or as probabilities, as appropriate) | *Parameter uncertainty*: All main text figures (expect Fig 1) and Supplementary Information (S1) Figs B–E |
| | *Prediction uncertainty*: All predictions incorporate structural and parameter uncertainty, although assessment of past transmission and reporting is the main focus of this study | *Prediction uncertainty*: See Fig 8 in main text and M in this SI. |

Continued on next page

Table I – continued from previous page

| Principle | What has been done to satisfy the principle? | Where in the manuscript is this described? |
|---|---|---|
| **5. Testable model outcomes** | | |
| | Previous versions of this model have undergone validation exercises (data censoring) to examine the robustness of the predictive ability of the model [14, 18, 21]. Whilst this was not performed here, the model is an updated version of those that have undergone validation, with updates based on critical review of model fits as data for different regions or time periods is included. As more years' data become available in the future, the model outputs shown here will be able to be compared to reported active and passive case data to test model predictions. As the focus here is on retrospective analysis rather than prediction, only one basic future strategy is projected (continuation of current strategy), however model posteriors could be used to simulate strategies that were actually conducted after 2021, output predicted case reporting and compared to future human case data. | The main text and SI contains model predictions for EoT probability. Model posteriors and code are available at OSF (`https://osf.io/jtrs9/`) |

[26] World Health Organization. Control and surveillance of human African trypanosomiasis: report of a WHO expert committee. Technical Report 984, World Health Organization, 2013.
